# Direct effect of genetic ancestry on complex traits in a Mexican population

**DOI:** 10.1101/2025.09.09.25335237

**Authors:** Siqi Wang, Jaime Berumen, Alejandra Vergara-Lope, Paulina Baca, Elizabeth Barrera, Fernando Rivas, Diego Aguilar-Ramirez, Rory Collins, Jonathan R Emberson, Michael Hill, Michael E Goddard, Loic Yengo, Alexander Strudwick Young, Jesus Alegre-Díaz, Pablo Kuri-Morales, Roberto Tapia-Conyer, Jason Torres, Peter M Visscher

## Abstract

Human populations differ in disease prevalences and in average values of phenotypes, but the extent to which differences are caused by genetic or environmental factors is unknown for most complex traits. Comparing phenotypic means across populations is confounded by environmental differences and comparisons based on polygenic predictors can lead to biased inference^1,2^. Family-based analyses of genetically admixed individuals offer a powerful framework for disentangling the direct and associated effects of genetic ancestry on phenotypes. Here, we leverage genetic data from admixed adults in the Mexico City Prospective Study (MCPS)^3,4^ to estimate within-family ancestry effects^5^. We quantified associations between genetic ancestry and 15 complex traits among 52,583 unrelated individuals and in 39,714 relatives across 17,627 families. At the population level, relative to a European ancestry baseline, we estimate an effect of Indigenous American (IAM) ancestry of −1.98 standard deviations (*P* < 2×10^-16^) for height and a natural log-odds ratio (lnOR) of 1.73 (95% confidence interval [CI] 1.54-1.92) for type 2 diabetes (T2D, *P* < 2×10^-16^), and multiple associations with other traits and ancestries. We estimated a within-family direct effect of IAM of −1.51 standard deviations (*P* = 1.02×10^-8^) for height and lnOR of 5.13 (95% CI 2.48-7.78, *P* = 1.51×10^-4^) for risk of T2D. These direct effects are supported by between-ancestry differences in polygenic burden and evidence of selection at trait-associated loci. In contrast, we found no evidence for a direct effect of ancestry on educational attainment or other study traits despite large and significant associations at the population level, implying environmental causes or confounding. Overall, this study provides an experimental design to study between-ancestry genetic effects for complex traits and reports significant ancestry differences for height, T2D, and metabolic-related traits in a genetically diverse population from Mexico City.

## Introduction

Within species, multiple populations can differ in the average genetic and phenotypic values of complex traits due to the evolutionary forces of natural selection and drift or by artificial selection in the case of agricultural populations and dog breeds^6,7^. Differences in allele frequencies at loci affecting traits, along with environmental differences, can lead to differences in phenotypes between subpopulations. In humans, some observable mean phenotypic differences are known to be caused by genetic differences among groups of different genetic ancestries, for example, pigmentation^8^, drug metabolism^9^ and the prevalence of single-gene disorders^10,11^. (Throughout this paper we use the term “genetic ancestry” to refer to inherited variation from ancestral populations that differ in allele frequencies at many loci in the genome, which can be inferred from genetic data. We acknowledge that there is no consensus on terminology^12,13^ and that human populations are not genetically discrete.) Human populations differ in phenotype means for complex traits like height and in disease prevalence for complex diseases like diabetes, but we know very little about the relative contribution of genetic and environmental factors, and their interactions, to these differences.

In admixed populations, the genomes of individuals vary in their proportions of different founder ancestries and these proportions can be estimated with genetic markers^14^. Ancestry proportions can be correlated with trait values at the population level^15–17^, but these proportions can be correlated with environmental differences, implying that any observed association could be caused by both genetic and environmental factors. In gene-trait association studies, the gold-standard for inferring direct effects of genotypes on phenotypes is to perform a within-family association analysis, which conditions on parental genotypes^18,19^. Within-family genetic variation is due to random segregation of genetic material during meiosis, therefore associations between within-family genotype variation and phenotype are due only to causal effects of inherited alleles^20–22^. Here, we use the same underlying logic to estimate the differing effects of genome-wide ancestry from different populations, which we term ‘direct effects of ancestry’^5^ in analogy to ‘direct genetic effects’ as estimated in family-based association studies. We use genome-wide genotyping data from related and unrelated individuals from the Mexico City Prospective Study (MCPS)^3,4^, including more than 17,000 families, to estimate direct ancestry effects on 15 complex phenotypes, including anthropometric traits, biomarkers, type 2 diabetes (T2D), and educational attainment (EA). An overview of the study design and analysis strategy is provided in Fig. 1.

**Fig. 1.**
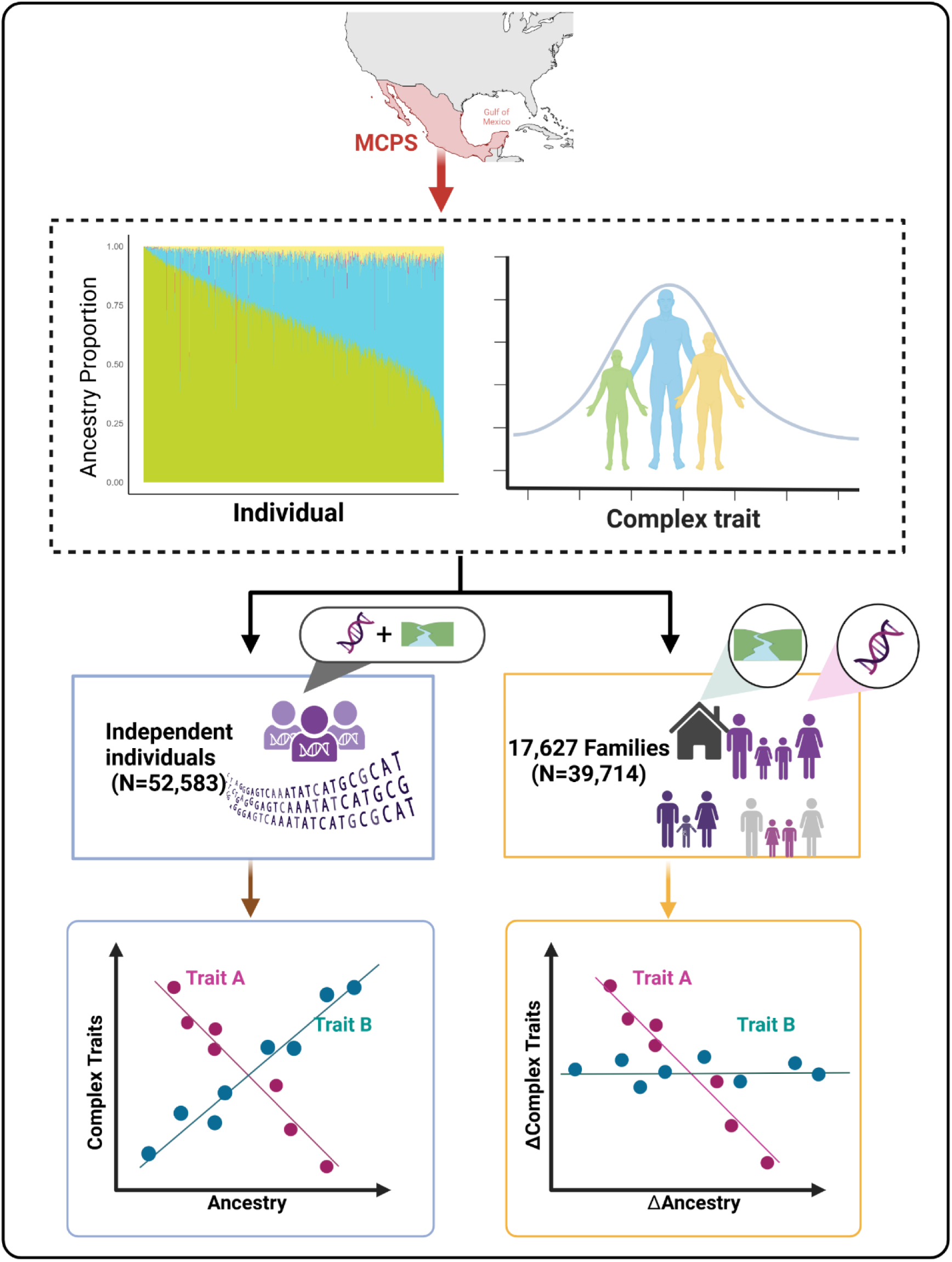
Graphical overview of the study. The aim of the study is to estimate direct and non-direct associated ancestry effects on complex traits using samples from an admixed populations from Mexico City (MCPS). Phenotypic trait values and genome-wide ancestry proportions vary between individuals in the population and within families. Ancestry-trait associations were estimated in two subsets: a sample of *n* = 52,583 unrelated individuals and a sample of *n* = 39,714 individuals from 17,627 families. Comparing the two subsets allows the separation of direct genetic effects and environmental effects that are associated with ancestry on traits. The same association in both sets is consistent with the association being driven by a direct genetic effect (Trait A). Conversely, the absence of an association within families is consistent with an environmental explanation (Trait B).

## Results

### Genetic ancestry is correlated between parents and segregates within families

Genome-wide proportions of Indigenous American (IAM), European (EUR), African (AFR) and East Asian (EAS) ancestries were derived for 140,829 individuals from MCPS^4^ (method). We formed two subsets: a sample of 52,583 individuals who were unrelated up to the fourth degree and a sample of 39,714 participants from 17,627 first-degree families (i.e. full-sibs and trios, see Methods) (Fig. 1, Extended Data Table 1). On average, genetic ancestry proportions were 67% (standard deviation (SD) 18%), 29% (SD 16%), 4% (SD 3%) and <1% (SD 2%) of IAM, EUR, AFR and EAS ancestry, respectively (Supplementary Figure 1, Extended Data Table 2). As expected, ancestry proportions were highly correlated with previously derived principal components (PCs) in MCPS^4^ (Supplementary Figure 2) as both principal components and admixture estimates are low-dimensional approximations of observed structure in genotype data^23^. The squared multiple correlation coefficient between ancestry proportion and PCs is up to 98% (Supplementary Table 1). From data on 2,847 families with both parents genotyped, we estimated the association between maternal and paternal genetic ancestry proportions and quantified the segregation variance within families. There is strong evidence for assortative mating on genetic ancestry, with correlations of 0.53, 0.52, 0.41 and 0.02 between estimated paternal and maternal ancestry proportions for IAM, EUR, AFR and EAS, respectively. Similarly, previously estimated PCs were positively correlated among parents (Supplementary Figure 3). Consistent with the evidence of substantial assortative mating on genetic ancestry, the variance of ancestry proportions between-families is much larger than the segregation variance. For IAM ancestry, the between-family SD is 0.167 whereas the within-family SD is only 0.020 (Extended Data Table 3). These results confirm that individuals in MCPS are highly admixed, that there is a positive correlation in estimated genome-wide ancestries between parents, and that genome-wide ancestry proportions segregate within families.

### Genetic ancestries associate with complex traits

We first quantified the association between genome-wide ancestry proportions and 15 complex traits in a sample of 52,583 unrelated individuals. For each trait, we regressed standardised trait values on genome-wide ancestry proportions and expressed estimated effect sizes (β, in SD units) relative to a 100% EUR baseline (Methods). Traits included height, weight, lipids and other biomarkers, type 2 diabetes and educational attainment (Methods, Extended Data Table 4).

Of the 45 trait-ancestry associations, 24 (13, 9 and 2 for IAM, AFR and EAS ancestry, respectively) were statistically significant at the study-wide significance *P* threshold of 5.89×10^-4^ (Methods), some with large effect sizes (Fig. 2, Supplementary Table 2). IAM ancestry was significantly associated with height (β = −1.98, *P* < 2×10^-16^), body mass index (BMI, β = 0.27, *P* < 2×10^-16^), low-density lipoprotein (LDL) cholesterol (β = −0.69, *P* < 2×10^-16^), apolipoprotein B (ApoB, β = −0.43, *P* < 2×10^-16^), T2D (natural log-odds ratio [lnOR] = 1.73 [95% CI 1.54-1.92], *P* < 2×10^-16^) and EA (β = −2.03, *P* < 2×10^-16^). We also considered a stricter definition of controls for T2D and a more fine-grained measure of EA (Methods). These sensitivity analyses showed a moderate increase in the effect of IAM on T2D risk and a small increase in the effect of IAM on EA (Supplementary Table 2). Genome-wide AFR ancestry proportions were also associated with several traits, including HbA1c (β = 0.92, *P* = 1.7×10^-8^), SBP (β = 0.91, *P* = 9.6×10^-9^) and EA (β = −3.24, *P* < 2×10^-16^). Relative to the EUR baseline, the IAM-associated effect sizes correspond to a reduction of approximately 13 cm in height and an increase in T2D prevalence of approximately 19%.

**Fig. 2:**
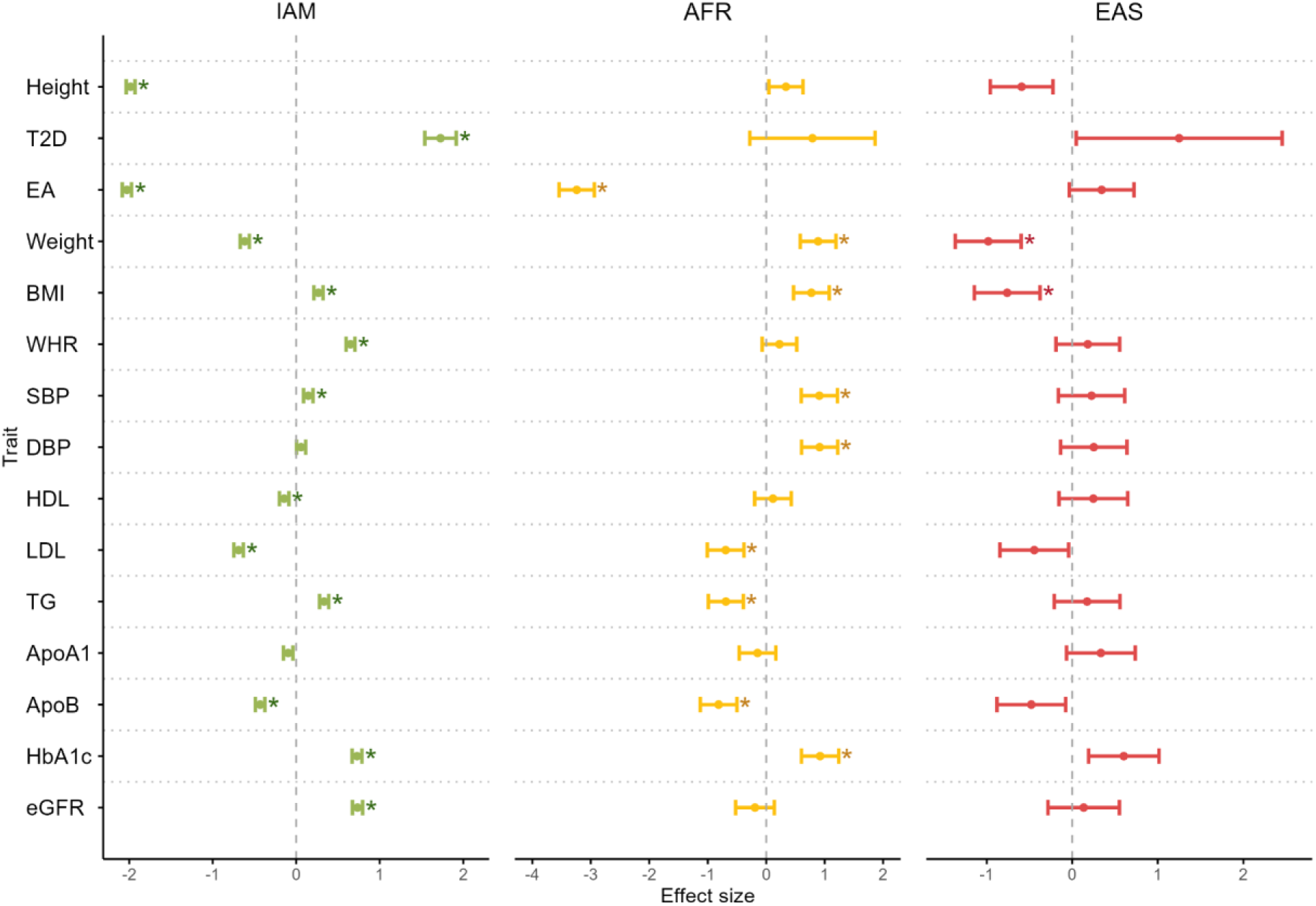
Association between ancestry and study traits in the MCPS. Analyses were conducted in a sample of unrelated individuals, with sample sizes ranging from 40,744 to 52,558 (Extended Data Table 4). Effect sizes are reported as lnOR for T2D and in standard deviation units for other phenotypes. Error bars represent 95% confidence interval. *Study-wide significant (*P* < 5.89×10^-4^). IAM = Indigenous American ancestry; AFR = African; EAS = East Asian; T2D = type 2 diabetes; EA = educational attainment; BMI = body mass index; WHR = waist-to-hip ratio; SBP = systolic blood pressure; DBP = diastolic blood pressure; HDL = high density lipoprotein; LDL = low density lipoprotein; TG = total triglycerides; ApoA1 = apolipoprotein A1; ApoB = apolipoprotein B; HbA1c = glycated haemoglobin; eGFR = estimated glomerular filtration rate.

Taken together, these results show that genome-wide ancestry proportions are strongly associated with anthropometric traits, biomarkers, T2D and educational attainment in MCPS, and that IAM ancestry is associated with increased T2D and its risk factors. However, these associations do not distinguish between direct ancestry effects and environmental effects that are correlated with genetic ancestry proportions. We therefore turned to the family data, where population-level associations can be partitioned into direct ancestry effects and between-family ancestry associations (Supplementary Note 1), and applied an analytical approach based on Mendelian segregation.

### Genetic ancestry associates with height and T2D within families

We utilized 17,627 families from MCPS, comprising 30,407 sibling pairs, simultaneously estimating both between- and within-family effects of genetic ancestry, using linear mixed models to take account of the covariance structure of the data (Methods; Supplementary Note 1). Genome-wide identity-by-descent (IBD) proportions were estimated between all sibling pairs (Methods) and were uncorrelated with pairwise estimated genetic ancestry (Extended Data Fig. 1). For families with no parental genotypes, we estimated the average parental ancestry proportions from the mean genetic ancestry of the genotyped offspring (Supplementary Figure 4, Supplementary Note 1).

For the 90 trait-ancestry combinations, we found three significant associations within families (Supplementary Table 3). There was a within family direct ancestry effect of IAM ancestry on height −1.51 SD (95% CI: [−2.02, −1.00], *P* = 1×10^-8^), whereas the between-family ancestry effect was not statistically significant (Fig. 3b), consistent with the population-level associations being driven by genetic effects, which are accounted for by the within-family effect. Similarly, we found a strong direct ancestry effect of IAM on T2D risk (lnOR = 5.13 [95% CI 2.48-7.78], *P* = 1.51×10^-4^). In contrast, the estimated direct ancestry effect for educational attainment was close to zero, despite a large and significant (β = −1.11, *P* = 2.51×10^-5^) between-family association (Fig. 3f). The associations of ancestries with all study traits are shown in Fig. 4a.

**Fig. 3:**
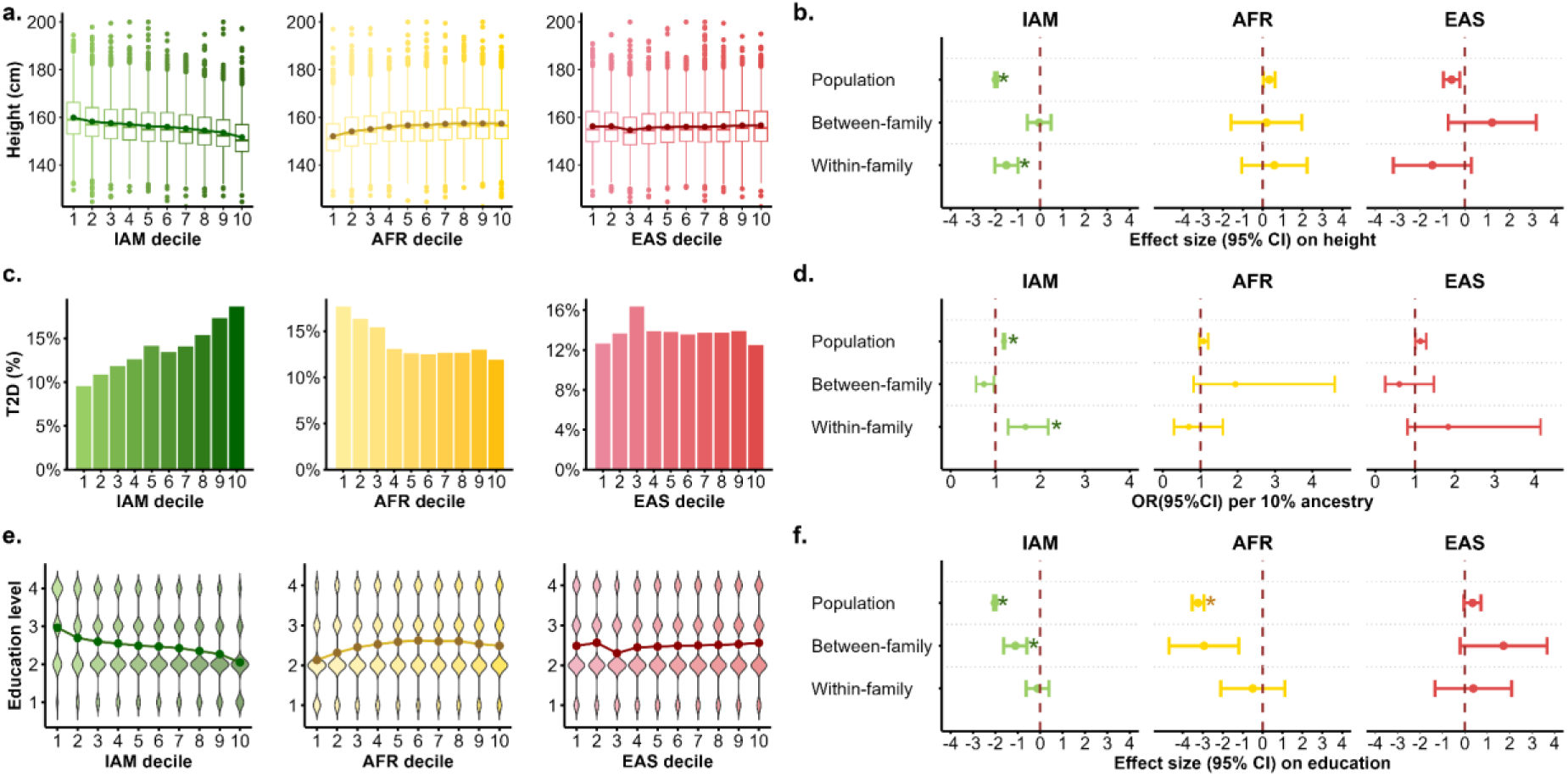
Between and within-family effects of ancestry on height, T2D and educational attainment. a, Height distribution across bins defined by ancestry proportion deciles. The dot with value represents the mean of height, the boxplot represents the median, interquartile range, and overall spread of the heights within each ancestry decile. b, Estimated ancestry effect for height. c, T2D prevalence by ancestry deciles in the population (*n* = 46,992). d, Effect size in odds ratio (OR) per ten percent ancestry on T2D^#^. e, Education level^##^ distribution by ancestry deciles in the population. f, Estimated ancestry effect for education level. Error bars represent 95% confidence interval. *Study-wide significant (*P* < 5.89×10^-4^). ^#^Between- and within-family effect sizes on T2D are from a generalised linear mixed model with a single random effect of family fitted (Methods). ^##^Education level (4=university or high school, 3=middle school, 2=elementary, 1=illiterate or literate). For each trait and each ancestry, population estimates are derived from unrelated individuals (*n* = 52,064 for height, *n* = 46,992 for T2D, *n* = 52,558 for education level). Between- and within-family effect sizes were estimated from family data (height, *n* = 39,298 individuals from 17,614 families; T2D, *n* = 35,385 individuals from 17,301 families; education level, *n* = 39,698 individuals from 17,627 families). Note that for panels a, c and e, the mean ancestry proportions in the deciles vary across ancestries. For IAM, AFR, and EAS, the mean ancestry proportions in the first and tenth deciles are (0.32, 0.95), (0.01, 0.11), and (10^-5^, 0.03), respectively.

**Fig. 4:**
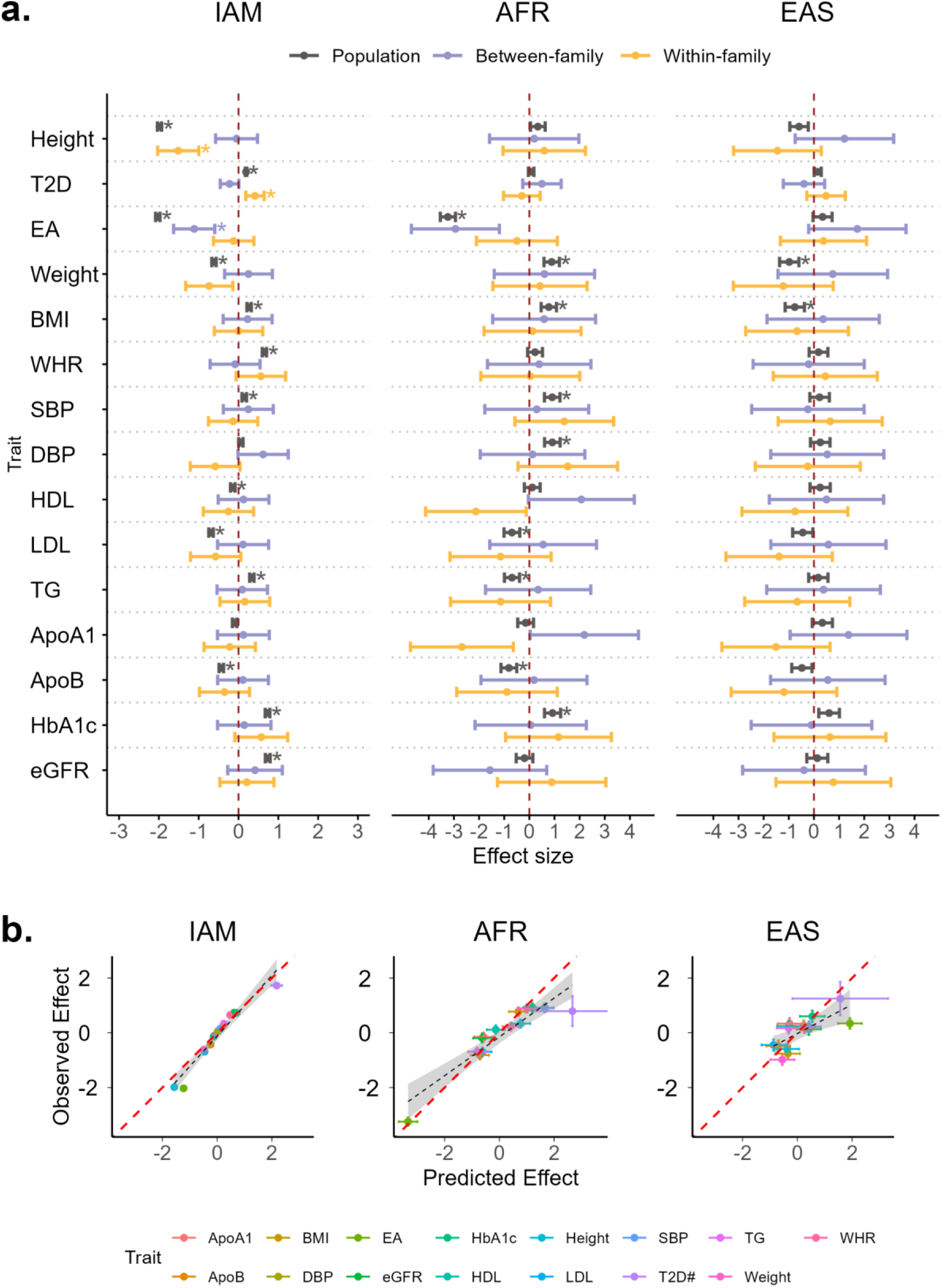
Between and within-family effects of ancestry on all 15 traits. a, Effect of different ancestries on all study traits. *Study-wide significant (*P* < 5.89×10^-4^, Methods). Error bars represent 95% confidence intervals. All population-level effect sizes shown in the figure were estimated from a linear model, and between- and within-family effect sizes were estimated by a mixed linear model, using the observed 0-1 scale for T2D. For each trait and each ancestry, population estimates are given from a sample of unrelated individuals, with sample sizes ranging from 40,744 to 52,558 (Extended Data Table 4). Between and within-family estimates are from family data, with sample sizes ranging from *n* = 30,500 (16,198 families) to *n* = 39,698 (17,627 families), Extended Data Table 4. b, Comparison between the ancestry effect sizes (beta ± SE) at the population level from the sample of unrelated individuals (y-axis, “observed”) and their estimates (beta ± SE) predicted (Methods) from the analysis of the family data (x-axis, “predicted”). #For T2D, effect sizes are in lnOR units, with the effect sizes from the family data estimated from a generalised linear mixed model with a single random effect of family (Methods).

As a by-product of the family analyses, we obtain estimates of within-ancestry genetic segregation variance and estimates of variance due to effects that are common to siblings but not explained by IBD sharing^24,25^. Estimates of heritability and the proportion of variance due to common environmental factors are shown in Extended Data Fig. 2 (Supplementary Table 4). Although the standard errors are large, estimates of heritability are substantial and much larger than estimates of shared environmental variance for nearly all traits. An exception is EA, where the data are consistent with all covariance among siblings being due to shared environmental factors. The more fine-grained measure of EA gave similar results (Supplementary Table 3). Given the large SE of the estimate, the apparent lack of genetic variation could be due to sampling error. Moreover, in these analyses, genetic variance in the population due to assortative mating would appear as common environmental variance^25–27^.

The estimates obtained from the family data predict the expected effect size from a population sample without family data (Supplementary Note 1). Fig. 4b shows strong concordance across all traits when comparing the effect sizes from the independent population and family samples.

In summary, among the 15 complex traits studied with the family data, we report evidence that ancestry has a direct effect on height and risk of T2D and a non-direct associated effect on educational attainment.

### Height and T2D polygenic loads are associated with genetic ancestry

The direct effect of ancestry on height and T2D is consistent with a substantial genetic contribution but uninformative with respect to genetic architecture. To investigate whether the estimated direct effects could be partly due to ancestry differences in the frequency of common trait-associated variants, we estimated a polygenic burden for each individual in the population sample from the largest multi-ancestry GWAS to date for height^28^ and T2D^29^ and correlated it with genome-wide ancestry proportions (Methods). For height, we found a large negative partial correlation between genome-wide IAM ancestry and polygenic load (r = −0.46, *P* < 1×10^-300^). Significant but small partial correlations were observed for AFR (r = −0.10, *P* = 1.17×10^-123^), and EAS (r = −0.06, *P* = 7.27×10^-40^). For T2D risk, there was a positive partial correlation between IAM ancestry and polygenic risk burden (r = 0.35, *P* < 1×10^-300^) and positive but smaller correlations for AFR (𝑟=0.12, *P* = 2.58×10^-176^) and EAS (r = 0.05, *P* = 9.61×10^-35^). A population specific polygenic load from common variants is unlikely to be an unbiased reflection of the true mean genetic value of that population as lead SNPs from GWAS are not necessarily causal variants and linkage disequilibrium (LD) patterns vary by population. As coding variants are more likely to be causal, we considered a burden of missense and loss-of-function variants, identifying 314 for height and 36 for T2D (Methods). Using the coding variant burden attenuated the association with genetic ancestry for height and gave inconclusive effects for T2D (Supplementary Table 5), likely due to a combination of a small number of variants and rare variant ascertainment bias, since most of the 36 missense variants have low minor allele frequency in Europeans (Supplementary Table 6), where most of the GWAS discovery data were ascertained.

If the polygenic burden captures a proportion of the direct ancestry effect we estimated in the family data, then adjusting for it should attenuate their estimated effect sizes. We therefore estimated the effects of ancestry and polygenic burden simultaneously in the family analysis and observed a significant attenuation (Supplementary Table 7). The IAM direct effect on height changed from −1.51 standard deviations to −0.43 (*P* > 0.05) and the IAM effect (lnOR) on T2D from 5.13 to 3.37 (*P* = 0.01). A smaller effect of IAM on height but no change for T2D was observed when the missense polygenic load was included (Supplementary Table 7).

These findings show that the direct ancestry effects for height and risk of T2D are consistent with differences in polygenic loads between ancestries involving common variants that are segregating across populations.

### Selection on height and T2D associated loci

We investigated whether the genetic differences between IAM and EUR ancestries for height and T2D could have been driven by natural selection. We obtained IAM allele frequencies by selecting 1,012 MCPS individuals whose estimated IAM ancestry in the genome was larger than 99.5% and validated those against frequencies from Indigenous Americans in the Human Genome Diversity Project (HGDP) (Methods, Supplementary Figure 5). We first estimated the mean fixation index (*F*_ST_) at the trait-associated loci between IAM and EUR ancestry populations for height and T2D and compared the estimates with control SNPs (Methods). Mean *F*_ST_ of associated loci was significantly higher than that of matched control SNPs for height (0.120 versus 0.116, *P* = 2.16×10^-4^), but not for T2D (0.110 versus 0.109, *P* = 0.83) (Supplementary Figure 6). We subsequently calculated the IAM-EUR difference in frequency of trait-increasing alleles at loci that were statistically associated with height and T2D in the

GWAS discovery sets^28,29^ and compared those to frequency differences of trait-increasing allele at control SNPs in the GWAS that were matched on frequency of trait-increasing alleles (fTIA) and LD score (Methods). The top 20 loci with the largest difference in fTIA between IAM and EUR for height and T2D are shown in Supplementary Table 8. The largest difference in fTIA is −0.87 for height (rs174570) and 0.75 for T2D (rs12625671). Fig. 5 shows that for both traits there is a systematic and significant difference in trait-increasing allele frequency, in the direction consistent with IAM having a lower burden of height-increasing alleles (*P* = 2.16×10^-5^) and a higher burden of T2D-increasing alleles (*P* = 1.48×10^-2^). Our test for selection is conservative because our control SNPs can also be associated with the trait even when not in LD with genome-wide significant loci, in particular for height where a randomly chosen SNP in the discovery data is genome-wide significant^28,30^. This is the likely explanation that the mean frequency difference of the null distribution is not zero and in the direction of the genetic mean difference (Fig. 5). As an additional control, we performed the same analyses on two traits, SBP and BMI, with well-powered GWAS summary statistics^31,32^ for which there is no evidence of a genetic difference between IAM and EUR ancestries. The results show no systematic difference in fTIA for these traits (Supplementary Figure 7).

**Fig. 5:**
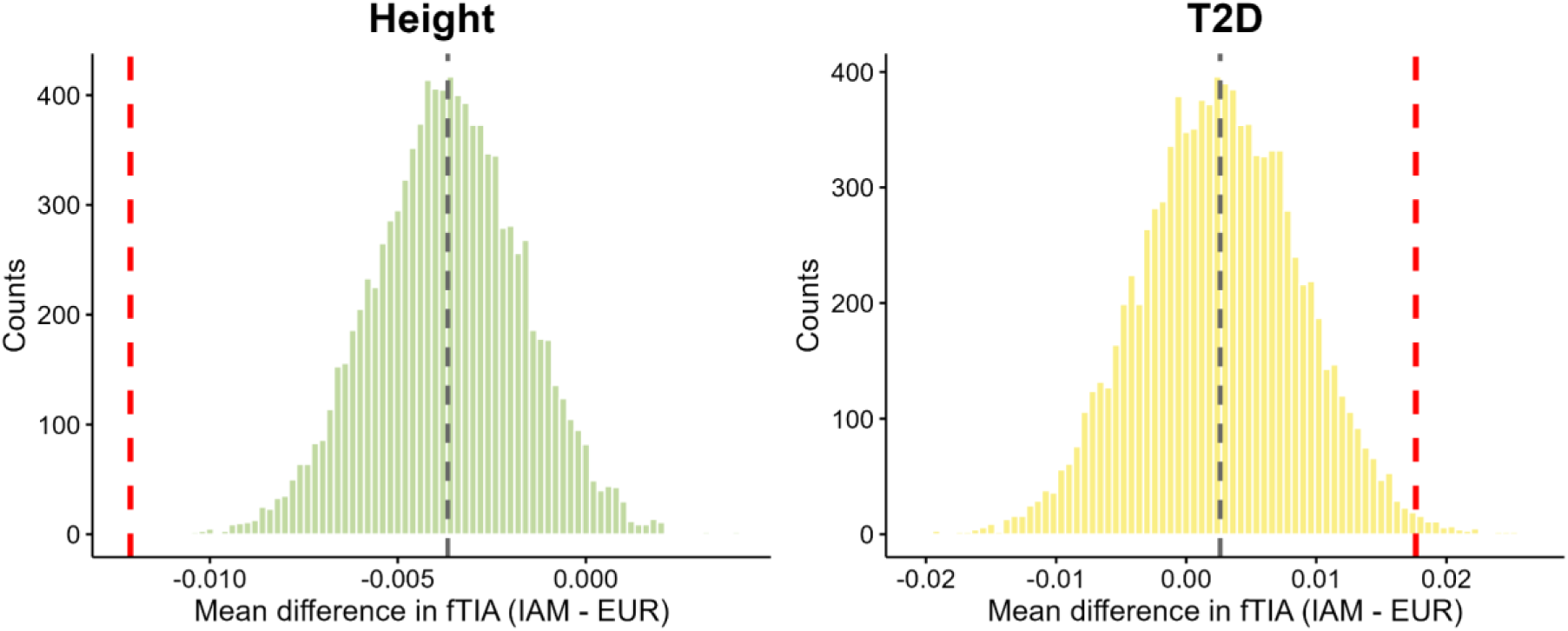
Mean difference in the frequency of trait-increasing alleles between IAM and EUR populations for height and type 2 diabetes. The histogram shows the distribution of fTIA differences for control SNPs, and the grey dashed line represents its mean value. The red dashed line indicates the mean value of the fTIA difference for the trait-associated SNPs. Control SNPs were matched on frequency and linkage disequilibrium scores of the genome-wide significant loci and repeatedly sampled from the GWAS summary statistics (Methods).

If genetic differences are partially due to selection, then loci that explained more trait variation when selection occurred should be more differentiated in frequency. We therefore correlated the difference in trait-increasing allele frequencies with their predicted amount of variance explained in Europeans (Methods), but did not detect a statistically significant association (Supplementary Figure 8).

The results from these analyses imply that the genetic difference between IAM and EUR ancestries in polygenic burden for height is partially driven by natural selection acting upon height-associated variants, while the evidence for selection on T2D-associated variants is inconclusive.

## Discussion

We used the natural experiment of randomization of genetic material during meiosis to estimate the causal effect of genetic ancestry differences on complex traits in an admixed sample from Mexico City. In principle this design can be applied whenever there is phenotypic and genome-wide genetic data from admixed families, enabling the disentanglement of causal effects of ancestry differences from correlations between genetic ancestry and phenotype due to other factors. By conducting a large family analysis in an admixed population, we have provided evidence that between-ancestry differences for complex traits can be genetic and explained by genome-wide differences in the frequency of trait-increasing (decreasing) alleles. Specifically, we found strong evidence for an IAM-EUR genetic difference in height and T2D risk in the MCPS, consistent with the direction of phenotypic differences and polygenic load between these ancestries.

While our within-family direct ancestry effect estimates can be given a causal interpretation, they do not imply a specific mechanism. It is possible that ancestry differences cause phenotype differences through social mechanisms such as ancestry-related appearance-based discrimination. However, our estimates are consistent with ancestry associated phenotype differences in height and T2D risk being entirely explained by direct ancestry effects (estimated within-family): if appearance related discrimination exerted an environmental effect on these phenotypes, it is likely this would also operate between families and induce a between-family ancestry association in our design, which we do not observe for height and T2D. When we fit educational attainment as a proxy for socio-environmental effects in the association model for height and T2D in the sample of independent individuals, we observe that higher education is associated with reduced risk of T2D and increased height (Supplementary Table 2). However, IAM is still strongly associated with both T2D and height, with slightly attenuated effect sizes after adjustment for educational attainment.

At the population level, genetic ancestry effects were large and statistically significant for most traits, yet some of these estimates did not carry over when estimating their effects within families. The difference was particularly striking for educational attainment where extremely large associations with ancestry (of 2 SDs and more) were estimated at the population level. However, our family-based analysis suggests that environmental factors correlated with ancestry rather than causal effects of alleles that differ in frequency between IAM and EUR ancestries explain the association between genetic ancestry and educational attainment in the MCPS. This shows that associations between genome-wide ancestry and phenotypes — especially socio-economic phenotypes — in the absence of family data could lead to misleading inference^33^. In addition to no evidence for a direct genetic ancestry effect on educational attainment, we found no evidence for any within-family genetic segregation variance in MCPS (Extended Data Fig. 2). However, the SE of the estimate of heritability is large (0.11) and there is evidence for direct genetic variation associated with educational attainment in populations of European ancestry^34,35^.

It is not known if our findings translate to phenotypic differences between non-admixed IAM and EUR populations. Our inference is strictly on MCPS participants from Mexico City, and our estimates are weighted towards offspring of parents with more ancestry differences between their paternal and maternal genomes, analogous to how family-based genetic association estimates are weighted towards families whose parents have greater levels of heterozygosity^36^. This implies that direct ancestry effect estimates from admixed families may differ from the genetic contribution to phenotype differences between more homogeneous samples. Systematic differences could be induced by assortative mating with respect to both phenotype and ancestry and if effects of alleles depend on genetic and environmental background (GxE and GxG interactions). Furthermore, environmental effects may differ in un-admixed populations compared to the admixed samples used here.

Lack of concordance of estimated effect sizes within families compared to those estimated in the population sample can also be due to reduced statistical power. At the population level, the precision of estimated effect sizes is proportional to the product of sample size *n* and the between-family variance in ancestry proportion v(π_b_), whereas it is approximately proportional to the product of the number of sibling pairs (*n_s_*) and the within-family variance v(π_w_) for the estimated effect size within families. For IAM ancestry, the ratios *n_s_*/*n* and v(π_w_)/v(π_b_) are 0.58 and 0.014, respectively (Extended Data Tables 1 and 3), implying more than a 100-fold difference in precision. Precision is also lower for binary traits compared to quantitative traits, as shown in our results for T2D. Although the estimate of the within-family effect of IAM ancestry is statistically significant, the estimate is extremely large and imprecise (the 95% CI of the lnOR is 2.48 to 7.78) and its sampling correlation with the between-family IAM effect is −0.99 (Supplementary Table 9). Therefore, even larger sample sizes are needed to separate within-family and between-family ancestry effects in admixed populations for common diseases.

In MCPS we found strong evidence that IAM ancestry has a direct effect on the risk of T2D and is enriched for common T2D risk alleles. This is, to our knowledge, the first causally identified evidence of a genetic difference in susceptibility to a common disease between ancestries that is associated with a difference in disease prevalence. There is a high prevalence of T2D in Mexico^37,38^, and diabetes is the overall leading cause of premature death in Mexico^39^. Our results are strongly supportive of a direct, ancestry-related genetic contribution to T2D burden in this population, beyond environmental influences such as a diabetogenic diet and limited healthcare access. Notably, the association between T2D burden and IAM ancestry does not appear to be explained by environmental factors shared within families.

We provided evidence that natural selection has acted upon height and T2D associated loci, in a direction consistent with our within-family estimates. These analyses are not informative about where and when selection took place nor its mechanism. Evidence of selection for reduced height has been reported in Peruvian individuals^40^, from the observation that a trait-decreasing missense variant (rs200342067) was at much higher frequency (0.047) in Peru than in Europe (<0.001). The frequency of that variant is 2.27% in the MCPS sample of 1,012 individuals with >99.5% IAM ancestry and the estimated effect size is also large (−0.29 SD, or approximately 2 cm, *P* < 2 × 10^-16^ among the set of unrelated individuals). Our results are consistent with selection for reduced height being widespread in Central and South America. The study in Peruvian individuals also estimated the association between ancestry proportions and height at the population level and reported an Indigenous American ancestry effect of −14.75 cm relative to a European ancestry baseline, remarkably close to the estimate in MCPS of −2 SD, which is approximately −13 cm. Of note, rs174570 has the greatest fTIA difference for height between IAM and EUR in the present study (Supplementary Table 8). It lies within the *FADS2* gene, which encodes an enzyme involved in the production of long-chain polyunsaturated fatty acids^41^. Selection at the *FADS2* region shows opposite patterns in Greenlandic Inuit^42^ and Europeans, reflecting adaptation to marine versus plant-based diets, respectively^43^.

Our approach is similar to that of Lin et al.^5^, who estimated ancestry effects between and within families on 26 traits in 697 families with Inuit-European admixture in Greenland. The authors reported significant within-family ancestry differences for weight-associated phenotypes and, despite lack of power, concordance in effect sizes estimated at the population level and within families. Their estimate for height was −15 cm for Inuit ancestry at the population level and −3.5 cm (standard error 6.8 cm, statistically not significantly different from zero) estimated within families. Statistical significance for each trait was determined from a false discovery rate (FDR) approach for each parameter tested, which is less stringent than our approach.

Our study has a few limitations. We focus on a select number of traits from a single admixed population where the majority of admixture is between ancestral Mesoamerican and European populations, sampled from a single location (Mexico City). Additional data from IAM-EUR admixed populations sampled in different locations would show how general our conclusions are regarding the differing effects of Indigenous American and European ancestries. Additional data on more traits and diseases in other admixed populations would allow a fuller quantification of between-population mean genetic differences for complex traits. In summary, our study has shown empirically that genetic ancestry differences in humans can cause differences between complex traits and diseases, and that these can be estimated using data on admixed families. Using data from the MCPS, we inferred that Indigenous American ancestry causes reduced height and increased risk of type 2 diabetes compared to European ancestry and that these differences were shaped, at least in part, by natural selection.

## Methods

### Study population and ethics declaration

The MCPS is a prospective cohort of over 150,000 adult participants from Mexico City^3,4^. The baseline survey took place between April 14 1998 and September 28 2004 and focused on households in two urban districts—Coyoacán and Iztapalapa. Residents aged 35 or older were invited to participate in the study. Of the 112,333 households with eligible residents, at least one individual from 106,059 households participated^3,4^. Ethics approval was obtained from the Mexican Ministry of Health, the Mexican National Council for Science and Technology, and the University of Oxford, UK. All participants provided written informed consent.

### Admixture analysis and estimation of genome-wide ancestry proportions

Whole-genome sequence data from the 1000 Genomes Project (1KG) and the HGDP were downloaded and filtered to the set of autosomal variants present on the Illumina Global Screening Array v2 (GSAv2). These datasets were then merged with the MCPS GSAv2 array dataset with 140,829 participants (previously quality controlled as described in Ziyatdinov et al.^4^ with the adjustment of filtering for genotype missingness prior to filtering for individual missingness, allowing us to retain 2,318 more participants than the 138,511 individuals evaluated in that study). This resulted in a merged dataset of 485,043 unambiguous bi-allelic SNPs. 1KG and HGDP participants representing four global “superpopulations” were designated as reference samples and included 765, 727, 658, and 408 individuals of African, East Asian, European, and Indigenous American ancestry, respectively. This reference set was then supplemented with 1,000 randomly selected MCPS participants unrelated to the 4th degree, resulting in a total of 1,408 Indigenous American samples and 3,558 reference samples overall.

Ancestry-specific allele frequencies and per-individual ancestry proportions were estimated with ADMIXTURE^14^ v1.3.0. An admixture model was fit for K = 4 ancestral populations inferred among the set of reference individuals using an unsupervised procedure. Ancestry proportions for the remaining set of 139,829 MCPS participants were then estimated via projection with the -P option. Each of the estimated K ancestries was assigned to a global “superpopulation” based on averages within the reference samples (e.g. the ancestry with the highest average proportion among European reference samples was assigned to an inferred European ancestral population). Ancestry proportions from each of the four inferred ancestral populations were used in subsequent analyses.

### Selection of population and family samples

From the 140,829 participants we selected two non-overlapping subsets, a set of families (consisting of two or more genotyped siblings and 0, 1 or 2 genotyped parents, Extended Data Table 1) and a separate set of individuals who were not related up to the fourth degree of relatedness. Pair-wise relatedness for all samples was inferred with KING^44^, using the option - -ibdseg, as previously described^4^. From the 30,450 sibling pairs identified by KING, individuals identified as full siblings were grouped into family units. To ensure that all pairs within a family were full siblings according to KING’s estimation, 25 individuals were removed. Additionally, 4 individuals were excluded to ensure that each family had no more than two putative parents. The filtration resulted in a family dataset comprising 30,407 full-sibling pairs within 16,187 families (*n* = 38,274). To refine the family structure further, we incorporated parent-offspring relationships, identifying individuals with genotype data for both parents, resulting in 1,440 complete parent-offspring trios. Consequently, the family-based dataset comprised 39,714 individuals with either full siblings or two genotyped parents. The “population sample” was selected from the 63,130 individuals who were unrelated up to the fourth degree of relatedness, excluding family set individuals and their parents, leading to a sample set of 52,583. Restriction to this unrelated set was done to minimize confounding due to pedigree relatedness in estimating population-level effects.

### Estimation of identity-by-descent (IBD) sharing among siblings

For each identified family, genome-wide IBD was estimated with snipar (single-nucleotide imputation of parents)^45^ based on genotype array data. Snipar employs a hidden Markov model (HMM) to infer IBD segments shared between siblings, achieving near-theoretical accuracy and reducing IBD errors compared to KING^45^. When running snipar, genotyping array variants were filtered based on the following criteria: minor allele frequency < 5%, significant deviation from Hardy-Weinberg equilibrium (*P* < 1×10^-4^), or missingness > 1%. Additionally, a genotyping error probability threshold of 4.5×10^-4^ was applied.

A comparison of estimated genome-wide IBD proportions among sibling pairs using either genetic or physical genome length is shown in Supplementary Figure 9. For subsequent analyses, we used the realised relationships among sibling pairs estimated from snipar, with IBD defined as the genome-wide proportion of genetic map length (cM) shared.

### Phenotype selection

Fifteen complex traits data at baseline were explored in the analysis, including height, weight, BMI, WHR, SBP, DBP, HDL, LDL, total triglyceride, ApoA1, ApoB, HbA1c, eGFR, an ordinal categorical variable education attainment, and one disease T2D. eGFR was calculated using the formula of the Chronic Kidney Disease Epidemiology Collaboration (CKD-EPI) 2021, which considers age, sex, and creatine plasma concentration^46^. SBP and DBP were adjusted by adding 15 or 10 separately if the participants took antihypertension medications^47^. Educational attainment was represented by a category variable education (4=university or high

school, 3=middle school, 2=elementary, 1=illiterate or literate), which is treated as a continuous trait in the analysis. As a sensitivity analysis we also considered a more fine-grained measure of education using 14 categories (Supplementary Table 10). The inverse normal distribution function was used to derive category thresholds for the 14 categories from their cumulative probabilities, and the mean z-score for each category was calculated assuming an underlying normal distribution with multiple thresholds. This transformation placed the ordinal EA 14 categories on an underlying normal scale. The phenotypic correlation between the 1-4 and the 14-category z-score scale in the entire MCPS cohort was 0.94.

Individuals were considered to have T2D if they reported a prior diagnosis of diabetes (diagnosed age ≥ 35) or were taking anti-diabetic medication at baseline. Individuals who reported a diagnosis before age 35 and were on insulin were considered likely to have type 1 diabetes and were excluded from the T2D case set. Participants without a prior diabetes diagnosis or on anti-diabetic medication and with HbA1c < 6.0% were classified as normoglycemic controls. The American Diabetes Association uses a HbA1c threshold < 5.7%^48^, and therefore we used this threshold to select controls in a sensitivity analysis. Phenotype outliers were excluded if height (cm) < 120 or > 200, weight (kg) < 35 or > 250, BMI (kg/m^2^) < 15 or > 60, and WHR < 0.5 or > 1.5. To make the effect scale comparable across phenotypes, continuous traits were adjusted for age and age^2^ and standardised (mean 0, SD 1) within each sex, and outliers were further excluded according to mean ± 5SD. Further adjustments were applied for specific traits: BMI was residualised in the standardization for WHR, and fasting duration was residualised for HDL, LDL, TG, ApoA1, and ApoB.

### Multiple testing adjustment

To determine the statistical significance of estimated ancestry effects, we calculated the total number of independent tests and adjusted the *P* threshold accordingly. Each trait had three estimates (one from the population sample and two from the family sample) for each of the three ancestries, resulting in a total of nine comparisons per trait. Given that the 15 traits were correlated (Supplementary Table 11), we estimated the effective number of independent traits using the eigenvalues of the phenotypic correlation matrix^49^. The estimated number of independent traits was 9.43, leading to a study-wide significance threshold of 5.89×10^-4^ (= 0.05 / [9×9.43]).

### Estimation of ancestry effects

For the sample of unrelated individuals, linear regression analyses were conducted using R (version 4.3.2) to estimate the association between ancestries and complex traits. District (Iztapalapa or Coyoacán) was included as a covariate for all traits. Sex, age and age^2^ were included as covariates for T2D analyses. HbA1c and eGFR were analyzed exclusively in the subset of non-diabetic individuals—defined as those without a T2D diagnosis, not taking anti-diabetic medication, and with HbA1c < 6.5%^50^. Additionally, sensitivity analyses were performed, adjusting for the first seven genetic PCs or without fitting district as covariates (Supplementary Table 12). Note that only the first seven PCs were used as these PCs had normally distributed SNP loadings across the genome – a signature of population structure – as opposed to non-normally distributed loadings indicative of long-range LD^4,51^.

Quantitative traits in the family data were analysed using linear mixed models, fitting individual genetic effects and shared environmental effects as random effects and between- and within-family ancestry effects (IAM, AFR, EAS) as fixed effects (Supplementary Note 1). These analyses were performed in GCTA^52^ (version 1.94.3). The covariance structure between the phenotypes of siblings was modelled by fitting the estimated IBD relationship matrix and a matrix for shared environmental effects, as done previously^24,25^. These models simultaneously estimate between- and within-family ancestry effects and variance components for genetic and shared environmental effects.

### Analysis of type 2 diabetes

T2D was the only binary (0-1) trait and was analysed using both linear (mixed) models and generalised linear (mixed) models. The statistical software packages we used did not have an option for a generalised linear mixed model with multiple random effect and user-specified covariance structures. We did have that option for linear mixed models and used that for T2D so that on the 0-1 scale it is the same model as for the quantitative traits. For the population set, we first fitted a simple linear model using *lm* in R (version 4.3.2) and subsequently fitted a generalised linear model with a logit link function, using *glm* in R. For the family data we fitted a linear mixed model on the 0-1 scale using GCTA, as was done for the quantitative traits, and transformed the variance components to a liability scale (as implemented in GCTA), assuming a population prevalence of 15.54%, which is the prevalence in the entire MCPS population with T2D data (*n*=125,042). We also fitted a generalised linear mixed model using *glmer* in R. To facilitate model convergence, age was rescaled to have a mean of 0 and a standard deviation of 1. However, this implementation can only fit a single random effect with a simple covariance structure and therefore we fitted family as the only random effect.

#### Polygenic burden analysis

We estimated a polygenic burden for height and T2D for each person in the sample of 52,583 unrelated individuals. For each of these traits, we used results from the largest trans-ancestry genome-wide association analysis study (GWAS) to date. For height we took the set of 12,111 SNPs from a conditional and joint (COJO) SNPs-analysis^28^. For T2D, we took 1,289 independent genome-wide significant variants from Suzuki et al.^29^. Height COJO variants and T2D GWAS variants were matched to MCPS genomes using previously TOPMED-imputed genotypes^4^ and for each individual, the polygenic burden was calculated by adding up the number of trait-increasing alleles, using the *--score sum* function in PLINK^53^ (v1.9). The Ensembl Variant Effect Predictor^54^ tool was used to make annotations for the trait associated SNPs of 12,111 COJO variants from Yengo et al.^28^ and the 1,289 independent genome-wide significant variants from Suzuki et al.^29^. Loss-of-function (LoF) variants were defined as the variants with annotation frameshift, stop gained, stop lost, start lost, splice acceptor, or splice donor^55^. A total of 314 (2 of 316 missing in MCPS) missense and LoF variants were identified for height-associated SNPs, and 36 (5 of 41 missing in MCPS) missense variants for T2D. To evaluate the association between polygenic burden and a specific ancestry while controlling for the effects of the other two ancestries, we conducted a partial correlation analysis using the ppcor^56^ package in R. To estimate the effect of ancestry after accounting for a polygenic burden we fitted the latter as a fixed effect in the mixed linear model analysis of the family data. For the T2D analysis, the polygenic burden was scaled to have a mean of 0 and a standard deviation of 1.

### Selection analyses

We followed the approach of Guo et al.^30^. Allele frequencies for EUR ancestry were taken from the independent European population (*n* = 348,658) in the UK Biobank. Allele frequencies for IAM were calculated from a sample of 1,012 individuals in MCPS who had an estimated genome-wide proportion of IAM ancestry > 99.5%. Since both the GWAS for height and T2D were from predominantly EUR samples, we used European samples (*n* = 503) of the 1000 Genomes reference to calculate LD scores, applying the *--ld-score* and *--ld-score-adj* functions in GCTA^52^ with a window size of 1,000 kb. SNPs from the GWAS summary statistics^28,29^ were matched with those having EUR frequency, IAM frequency, and LD score, resulting in 1,183,522 and 8,447,340 variants for height and T2D, respectively. The fTIA in IAM and EUR was calculated for the trait-associated 12,111 (11,778 available) COJO variants from Yengo et al.^28^ and the 1,289 (1,208 available) genome-wide significant variants from Suzuki et al.^29^. *F*_ST_ for each of the associated loci was calculated from Hudson’s *F*_ST_ equation^57^ *F*_ST_ = [(p̃_1_ - p̃_2_)^2^ - (p̃_1_(1 - p̃_1_)/(n_1_ - 1)) - (p̃_2_(1 - p̃_2_)/(n_2_ - 1))] / [p̃_1_(1 - p̃_2_) + p̃_2_(1 - p̃_1_)], where p̃_1_ is the estimated fTIA in IAM, p̃_2_ the estimated fTIA in EUR, and n_i_ the sample size. To generate a null distribution, control SNPs matched on fTIA in EUR and LD score were sampled from the GWAS summary statistics (excluding the trait-associated variants), and their fTIA were calculated in IAM and EUR^30^. This was repeated 10,000 times, both for the analysis of *F*_ST_ and the analysis of the difference in trait-increasing alleles.

To correlate fTIA differences with effect sizes for the genome-wide trait-associated variants, we multiplied the square of the estimated effect size in the GWAS with expected heterozygosity (2×fTIA×(1-fTIA)) in EUR (Supplementary Figure 8).

For the association between SNP rs200342067 and height, we performed linear regression analyses in the set of 52,583 unrelated individuals, adjusting for ancestries, age, age^2^, sex, and district.

### Comparison of estimates from the population and family data

To explore concordance of the estimated ancestry effects from the family and population datasets, we used the estimated between and within-family effect sizes from the family data, predicted the population effect size (Supplementary Note 2) and then compared the predicted values with the “observed” estimate from the independent population sample. The predicted value is a linear combination of the between and within-family effect size and we derived its sampling variance (and standard error) using the *--reml-est-fix-varcov* function in GCTA, which provides the full variance-covariance matrix of the fixed effect estimates from the mixed model analysis.

## Acknowledgements

We thank the MCPS study participants, field workers, and research staff. Moreover, this work was supported by funding from the Mexican Health Ministry; the National Council for Science and Technology (CONACyT) for Mexico; the Wellcome Trust [058299/Z/99]; Cancer Research UK; the British Heart Foundation [RE/13/1/30181]; and the UK Medical Research Council [MC_UU_00017/2, MR/Z504543/1]. Genotyping was supported through an academic partnership involving the National Autonomous University of Mexico, the University of Oxford, Regeneron Pharmaceuticals, and AstraZeneca. No authors received payment from a pharmaceutical company or other agency for writing this article. This study also made use of data from the UK Biobank (project ID 12505). We thank Junren Hou for developing a new function to generate the full (co)variance matrix for GCTA REML analysis, Tian Lin for help with the GWAS summary statistics, and Julia Sidorenko for help with software.

## Author contributions

P.M.V. and J.T. conceptualized and jointly supervised the study. S.W. conducted statistical analyses, data visualization, and interpretation with supervision from P.M.V. and J.T. J.T. performed admixture analyses for the MCPS cohort. J.B., R.T.C., A.V.L., P.B., E.B., M.G., L.Y., A.S.Y., J.Y., F.R., D.A.R., R.C., J.R.E, M.H., J.A.D., P.K.M. contributed through suggestions and comments on methods, analyses, and result interpretation. R.T.C., J.B., J.T., A.V.L., P.B., E.B., F.R., D.A.R., R.C., J.R.E, M.H., J.A.D., P.K.M. contributed to data collection, preparation, management, and scientific leadership of the MCPS cohort. S.W., P.M.V., and J.T. wrote the manuscript with the participation of all authors. All authors reviewed and approved the final version of the manuscript.

## Rights retention

For the purpose of Open Access, the author has applied for a CC BY public copyright licence to any Author Accepted Manuscript (AAM) version arising from this submission.

## Data Availability

Data from the Mexico City Prospective Study are available to bona fide researchers. The study’s Data and Sample Sharing policy (https://www.ctsu.ox.ac.uk/research/mcps) can be viewed (in English or Spanish). The questions utilized in the study, along with the available study data, can be accessed and reviewed through the study’s Data Showcase (https://datashare.ndph.ox.ac.uk/mexico/).

## Code availability

The code used in this study is available from GitHub at https://github.com/mcps-analysts/Direct-effect-of-genetic-ancestry-on-complex-traits

## Competing interest declaration

The authors declare no competing interests.

## Additional information

Supplementary Note 1: Models to estimate ancestry effects between and within families.

Supplementary Table 1. Regression results of ancestry proportions with PC1–PC7.

Supplementary Table 2. Ancestry associations with complex traits at the population level.

Supplementary Table 3. Direct and indirect effects of ancestry on complex traits within families.

Supplementary Table 4. Variance components for all study traits.

Supplementary Table 5. Partial correlation of polygenic load with ancestries.

Supplementary Table 6. The 36 missense variants were identified from the 1289 independent single-nucleotide variants reported in Suzuki et al.’s multi-ancestry meta-regression on type 2 diabetes.

Supplementary Table 7. The effect of polygenic load on height and T2D.

Supplementary Table 8. Top 20 genetic loci showing the largest differences in trait-increasing allele frequencies between European and Indigenous American populations for height and type 2 diabetes.

Supplementary Table 9. The sampling variance–covariance and correlation matrix of the estimated fixed effects for T2D from two mixed models.

Supplementary Table 10. Categories and codes for educational attainment.

Supplementary Table 11. Pairwise Pearson correlation coefficients for traits.

Supplementary Table 12. Sensitivity analysis of ancestry-complex trait associations at the population level.

## EXTENDED DATA

**Extended Data Fig. 1.**
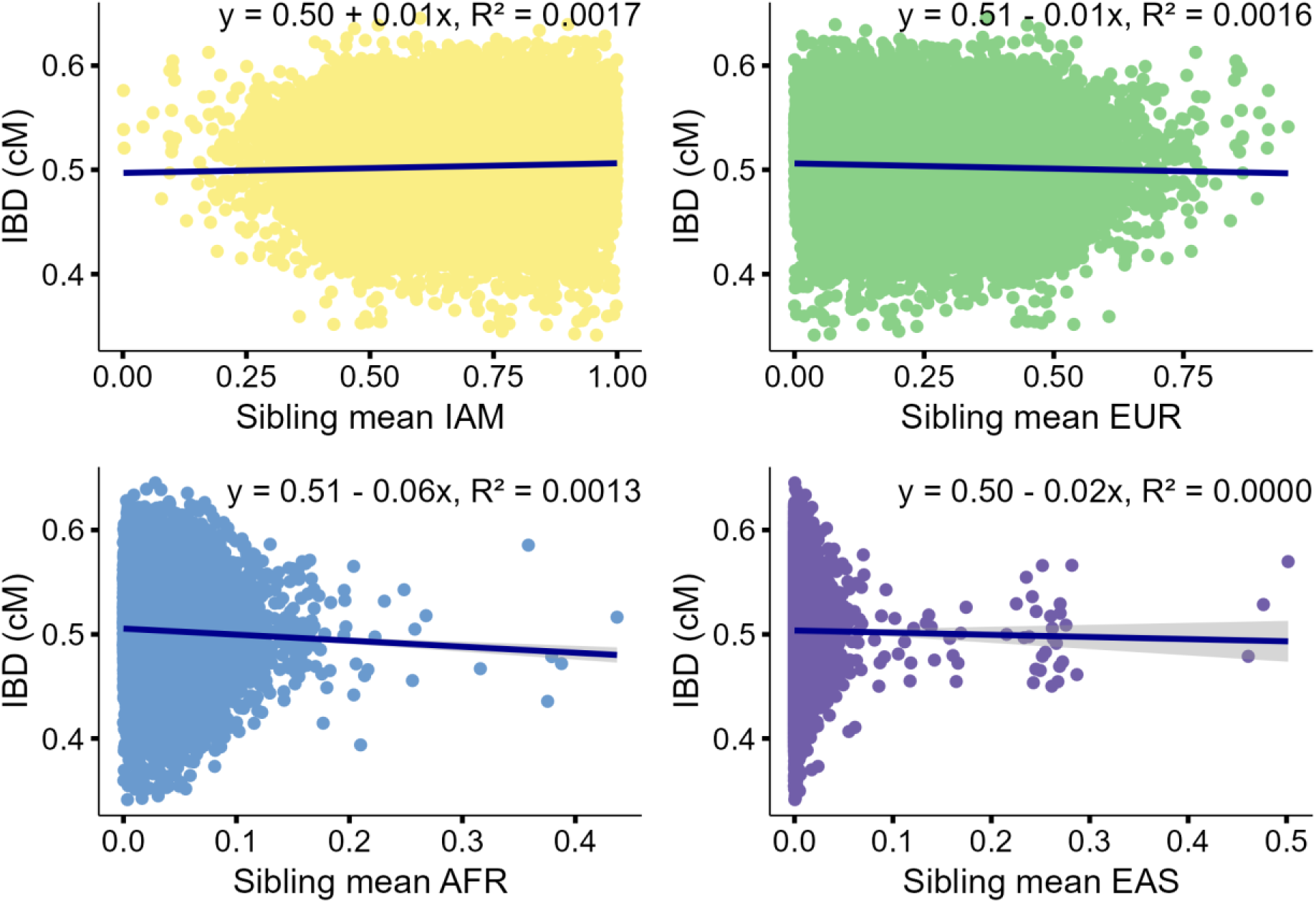
**Inferred genome-wide Identity-By-Descent (IBD) proportions are uncorrelated with sibling mean ancestry proportions.** Analyses were performed on *n* = 30,407 sibling pairs, using IBD estimated from snipar. Abbreviation: IBD = identical-by-descent, cM = centimorgan, IAM = Indigenous American, EUR = European, AFR = African, EAS = East Asian.

**Extended Data Fig. 2.**
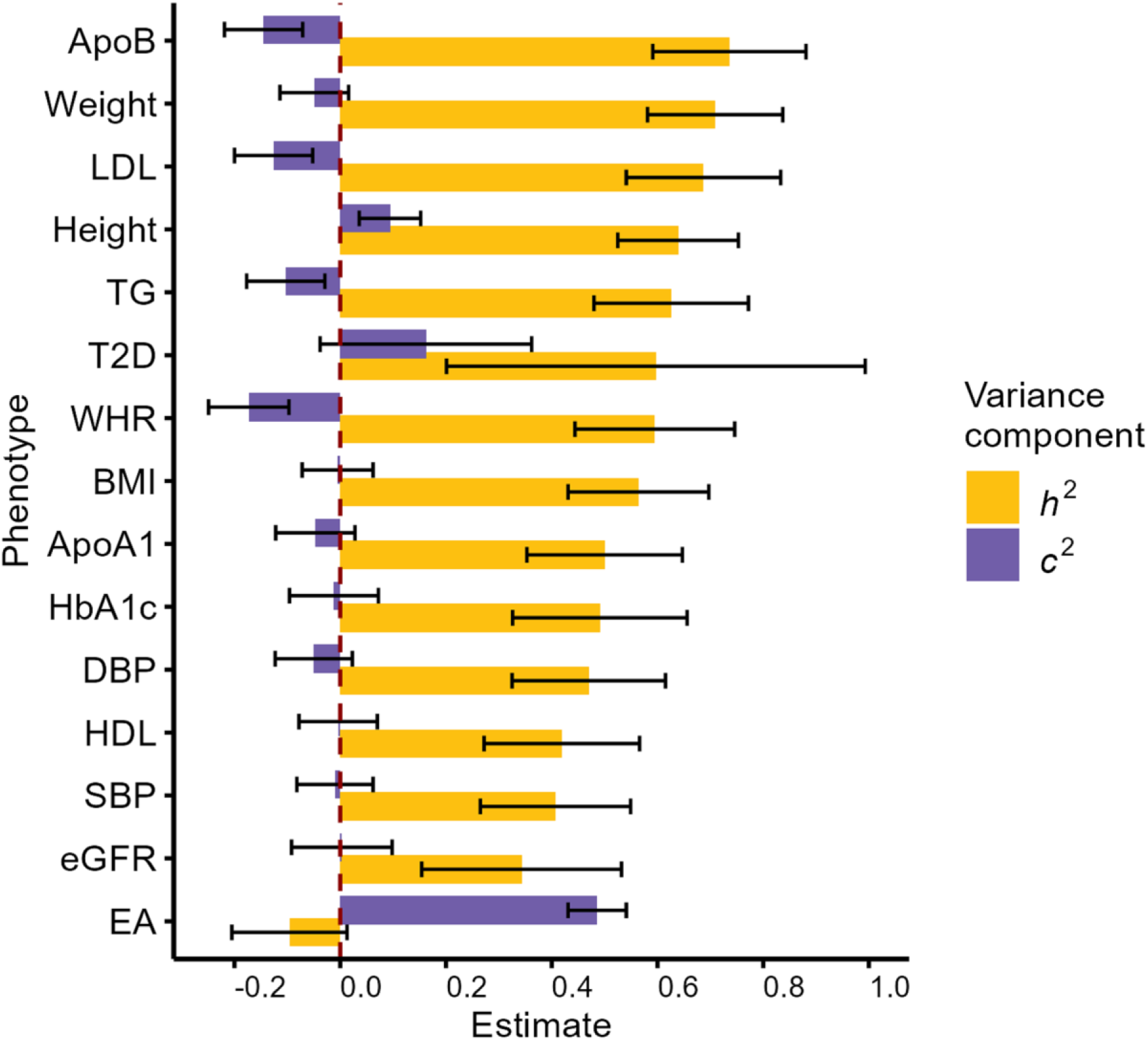
**Variance components estimation.** Bar plot showing estimates of the proportion of phenotypic variance explained by IBD sharing (*h^2^*) and other effects (*c^2^*). Error bars representing standard errors. Results are presented for T2D on the scale of liability. Abbreviation: ApoB = apolipoprotein B; LDL = low density lipoprotein; TG = total triglycerides; T2D = type 2 diabetes; WHR = waist-to-hip ratio; BMI = body mass index; ApoA1 = apolipoprotein A1; HbA1c = glycated haemoglobin; DBP = diastolic blood pressure; HDL = high density lipoprotein; SBP = systolic blood pressure; eGFR = estimated glomerular filtration rate; EA = educational attainment.

**Extended Data Fig. 3.**
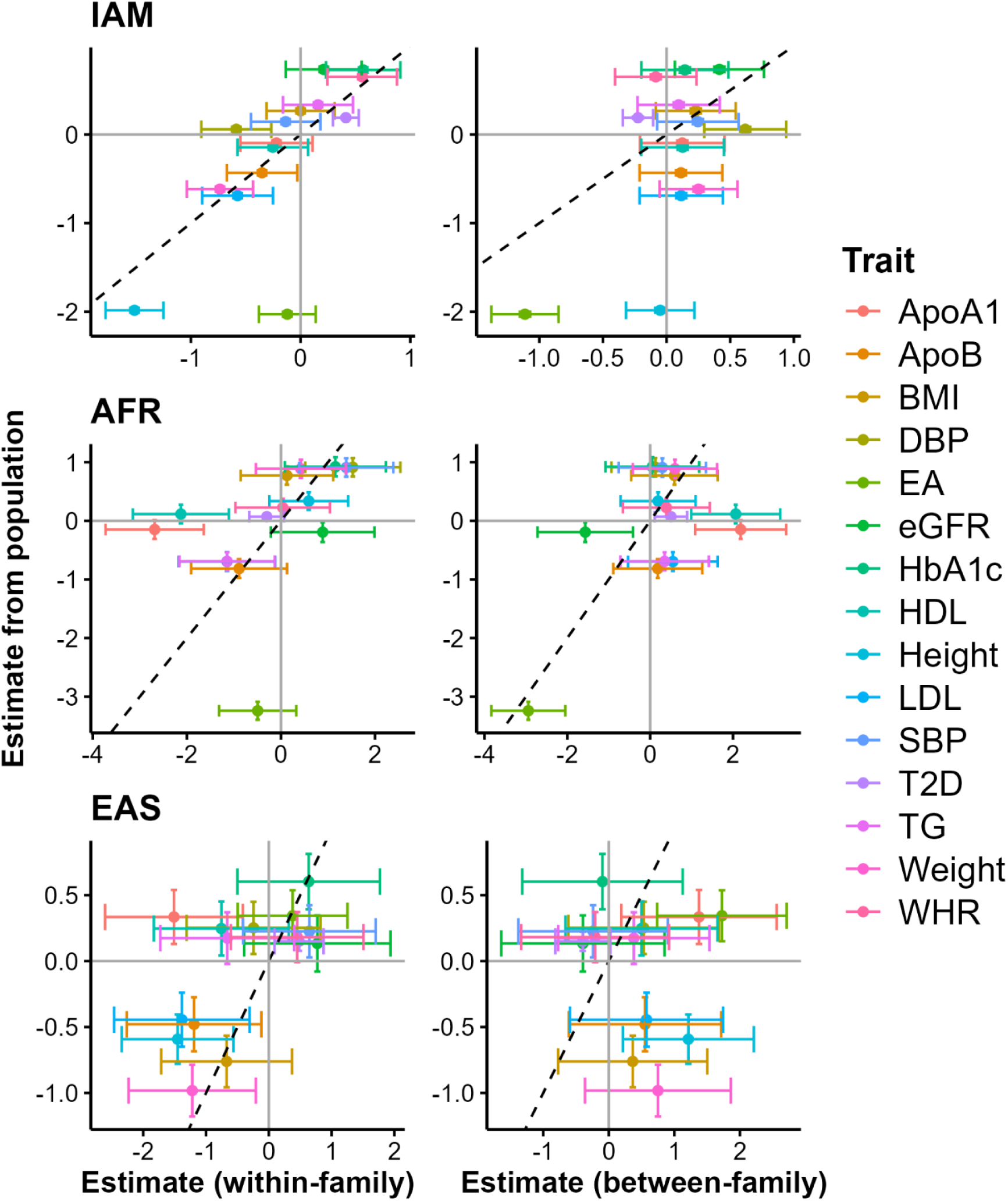
**Comparison of ancestry proportion effects on all study traits from population and family data.** All population-level effect sizes shown in the figure were estimated from a linear model, and between- and within-family effect sizes were estimated by a mixed linear model, using the observed 0-1 scale for T2D. Error bars represent standard errors. Abbreviation: IAM = Indigenous American ancestry; AFR = African; EAS = East Asian; ApoA1 = apolipoprotein A1; ApoB = apolipoprotein B; BMI = body mass index; DBP = diastolic blood pressure; EA = educational attainment; eGFR = estimated glomerular filtration rate; HbA1c = glycated haemoglobin; HDL = high density lipoprotein; LDL = low density lipoprotein; SBP = systolic blood pressure; T2D = type 2 diabetes; TG = total triglycerides; WHR = waist-to-hip ratio.

**Extended Data Table 1.**
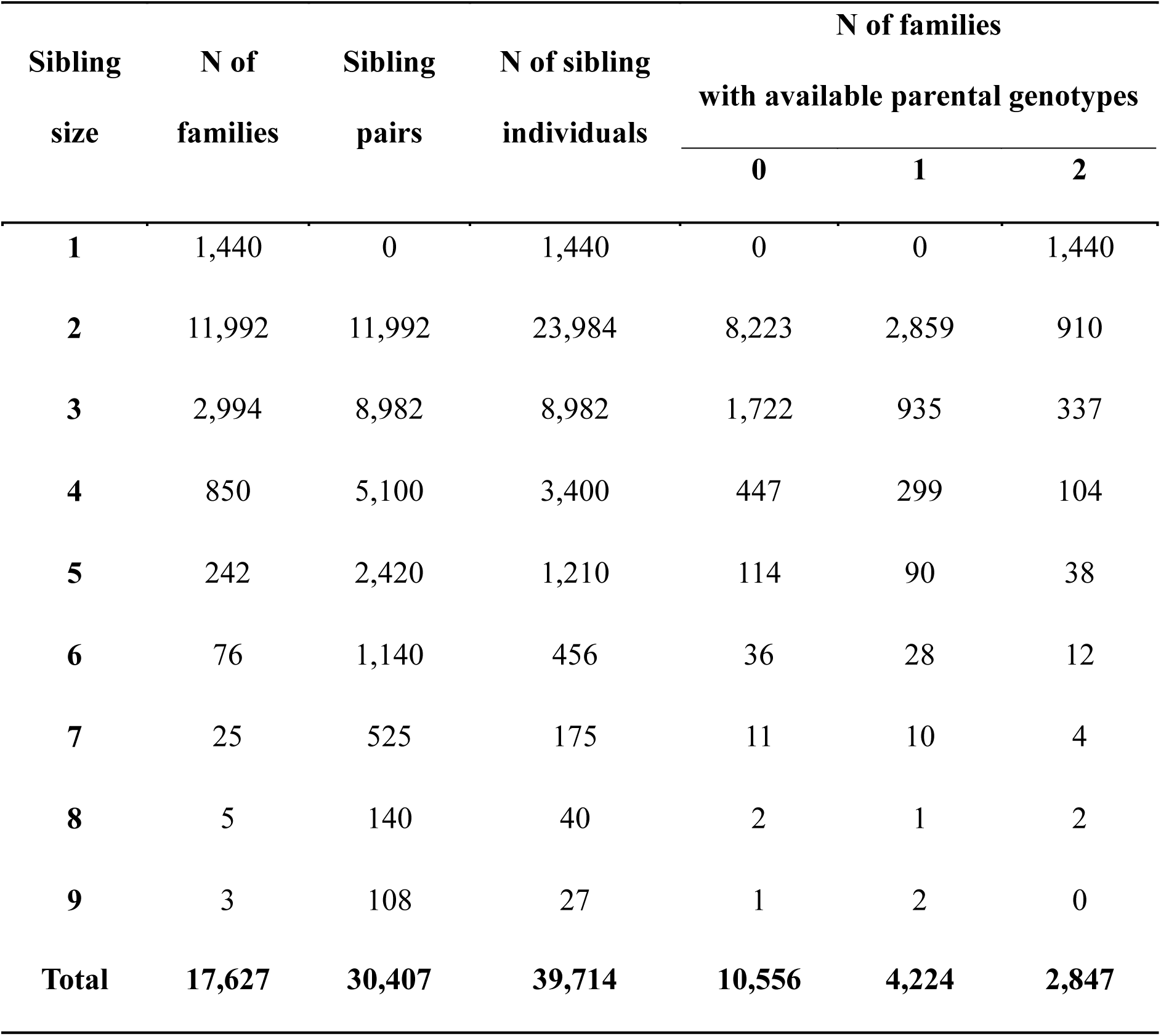
Distribution of family sizes.

| **Sibling**<br>**size** | **N of**<br>**families** | **Sibling**<br>**pairs** | **N of sibling**<br>**individuals** | **N of families**<br>**with available parental genotypes** |  |  |
| --- | --- | --- | --- | --- | --- | --- |
|  |  |  |  | **0** | **1** | **2** |
| **1** | 1,440 | 0 | 1,440 | 0 | 0 | 1,440 |
| **2** | 11,992 | 11,992 | 23,984 | 8,223 | 2,859 | 910 |
| **3** | 2,994 | 8,982 | 8,982 | 1,722 | 935 | 337 |
| **4** | 850 | 5,100 | 3,400 | 447 | 299 | 104 |
| **5** | 242 | 2,420 | 1,210 | 114 | 90 | 38 |
| **6** | 76 | 1,140 | 456 | 36 | 28 | 12 |
| **7** | 25 | 525 | 175 | 11 | 10 | 4 |
| **8** | 5 | 140 | 40 | 2 | 1 | 2 |
| **9** | 3 | 108 | 27 | 1 | 2 | 0 |
| **Total** | **17,627** | **30,407** | **39,714** | **10,556** | **4,224** | **2,847** |

**Extended Data Table 2.** Ancestry proportions (mean [SD]) of individuals, sibling pairs, families, and parent pairs.

| **Ancestry proportion** | **Individual level** |  |  | **Sibling mean**<br>**(N=30,407)** | **Family mean**<br>**(N=17,627)** | **Parental mean**<br>**(N=2,847)** |
| --- | --- | --- | --- | --- | --- | --- |
|  | All study population<br>(n=92,297) | Unrelated set<br>(n=52,583) | Family set<br>(n=39,714) |  |  |  |
| **IAM** | 0.67 (0.18) | 0.65 (0.18) | 0.71 (0.17) | 0.72 (0.17) | 0.70 (0.17) | 0.69 (0.16) |
| **EUR** | 0.29 (0.16) | 0.31 (0.17) | 0.26 (0.15) | 0.25 (0.15) | 0.26 (0.15) | 0.27 (0.15) |
| **AFR** | 0.04 (0.03) | 0.04 (0.03) | 0.03 (0.02) | 0.03 (0.02) | 0.03 (0.02) | 0.04 (0.02) |
| **EAS** | $3.87 \times 10^{-3}$ (0.02) | $4.33 \times 10^{-3}$ (0.02) | $3.25 \times 10^{-3}$ (0.01) | $3.11 \times 10^{-3}$ (0.01) | $3.31 \times 10^{-3}$ (0.01) | $3.75 \times 10^{-3}$ (0.02) |

**Extended Data Table 3.** Between-family and within-family variance of ancestry proportions.

| Variance | Parental and offspring genotypes* |  |  | Offspring genotypes only<sup>#</sup> |  |  |
| --- | --- | --- | --- | --- | --- | --- |
| | Between | Within | $\lambda$ | Between | Within | $\lambda$ |
|  | family | family |  | family | family |  |
| **IAM** | 2.61E-02 | 4.22E-04 | 0.016 | 2.79E-02 | 3.85E-04 | 0.014 |
| **EUR** | 2.13E-02 | 3.25E-04 | 0.015 | 2.28E-02 | 3.11E-04 | 0.014 |
| **AFR** | 4.75E-04 | 3.56E-05 | 0.075 | 5.41E-04 | 3.45E-05 | 0.064 |
| **EAS** | 2.33E-04 | 4.33E-05 | 0.186 | 1.35E-04 | 3.06E-05 | 0.227 |

\*Results show the estimates of the between and within-family variance of ancestry proportions. For the 2,847 families with parental genotypes, the between-family variance is the variance of the parental mean, and the within-family variance is the residual variance from a regression of ancestry proportion on the parental mean. <sup>#</sup>Using all 16,187 families with at least 2 genotyped offspring, the estimates of between and within-family variance are from a linear mixed model with family as a random effect. Parameter $\lambda$ is the ratio of the within to between variance (Supplementary Note 1). For the prediction results (Eq(2) in Supplementary Note 1), the values of $\lambda$ from the larger sample size were used.

**Extended Data Table 4.** Baseline characteristics of study population.

| Characteristic | All study population |  | Unrelated set |  | Family set |  | Family number |
| --- | --- | --- | --- | --- | --- | --- | --- |
|  | Mean (SD) | N (%) | Mean (SD) | N (%) | Mean (SD) | N (%) |  |
| **Age (years)** | 50.97(12.09) | 92,297 | 51.81 (12.36) | 52,583 | 49.85 (11.63) | 39,714 | 17,627 |
| **District (Iztapalapa)** | -- | 55,217 (59.83%) | -- | 30,040 (57.13%) | -- | 25,177 (63.40%) | 17,627 |
| **Male** | -- | 30,439 (32.98%) | -- | 16,545 (31.46%) | -- | 13,894 (34.99%) | 17,627 |
| **Height (cm)** | 156.07 (8.98) | 91,362 | 156.04 (9.04) | 52,064 | 156.12 (8.90) | 39,298 | 17,614 |
| **Weight (kg)** | 70.73 (13.38) | 91,457 | 70.50 (13.41) | 52,113 | 71.03 (13.34) | 39,344 | 17,616 |
| **BMI (kg/m<sup>2</sup>)** | 29.03 (4.98) | 91,346 | 28.95 (4.99) | 52,049 | 29.14 (4.98) | 39,297 | 17,615 |
| **WHR** | 0.90 (0.07) | 91,363 | 0.90 (0.07) | 52,067 | 0.90 (0.07) | 39,296 | 17,616 |
| **SBP\* (mm Hg)** | 129.30 (18.99) | 92,212 | 129.84 (19.30) | 52,538 | 128.59 (18.55) | 39,674 | 17,627 |
| **DBP\* (mm Hg)** | 84.60 (11.59) | 92,212 | 84.87 (11.72) | 52,538 | 84.24 (11.41) | 39,674 | 17,627 |
| **HDL (mmol/l)** | 1.00 (0.21) | 90,046 | 1.01 (0.22) | 51,307 | 1.00 (0.21) | 38,739 | 17,515 |
| **LDL (mmol/l)** | 2.47 (0.79) | 90,046 | 2.48 (0.79) | 51,307 | 2.45 (0.78) | 38,739 | 17,515 |
| **Total triglyceride (mmol/l)** | 1.57 (0.66) | 90,046 | 1.56 (0.65) | 51,307 | 1.58 (0.67) | 38,739 | 17,515 |
| **ApoA1 (g/l)** | 1.22 (0.19) | 90,046 | 1.23 (0.19) | 51,307 | 1.22 (0.19) | 38,739 | 17,515 |
| **ApoB (g/l)** | 0.90 (0.21) | 90,046 | 0.90 (0.21) | 51,307 | 0.89 (0.21) | 38,739 | 17,515 |
| **HbA1c (%)<sup>#</sup>** | 5.45 (0.37) | 75,707 | 5.45 (0.37) | 43,329 | 5.45 (0.37) | 32,378 | 16,628 |
| **eGFR (mL/min/1.73 m<sup>2</sup>)<sup>#</sup>** | 102.68 (14.68) | 71,244 | 101.66 (15.05) | 40,744 | 104.05 (14.06) | 30,500 | 16,198 |
| **Education level\*** | -- | 92,256 | -- | 52,558 | -- | 39,698 | 17,627 |
| **Level 1** | -- | 9,901 (10.73%) | -- | 6,191 (11.78%) | -- | 3,710 (9.35%) | -- |
| **Level 2** | -- | 43,169 (46.79%) | -- | 23,895 (45.46%) | -- | 19,274 (48.55%) | -- |
| **Level 3** | -- | 23,872 (25.88%) | -- | 13,062 (24.85%) | -- | 10,810 (27.23%) | -- |
| **Level 4** | -- | 15,314 (16.60%) | -- | 9,410 (17.90%) | -- | 5,904 (14.87%) | -- |
| **Type 2 diabetes** | -- | 11,669 (14.17%) | -- | 6,479 (13.79%) | -- | 5,190 (14.67%) | 17,301 |

Continuous variables are presented as mean ± SD and binary variables as number and percentage. SD=Standard deviation, N=Number, BMI=Body mass index,
WHR=Waist-hip ratio, SBP\*=Systolic blood pressure adjusted for hypertension medication, DBP\*=Diastolic blood pressure adjusted for hypertension medication,
HDL=High-density lipoprotein cholesterol, LDL=Low-density lipoprotein cholesterol. <sup>#</sup> Within non-diabetic population. \*Educational level (4=university or high school, 3=middle school, 2=elementary, 1=illiterate or literate).

## SUPPLEMENTARY INFORMATION

### Supplementary Methods

#### Supplementary Note 1: Models to estimate ancestry effects between and within families

This supplementary note provides a derivation of the expectation of regression coefficients to estimate between and within family ancestry effects on complex traits in family and population data and gives details of the models that were applied to the MCPS data. The objective is to clarify the interpretation of regression coefficients within the context of genetic ancestry studies in populations and families.

#### Notation

To keep the notation simple, we initially assume admixture of only two ancestries whose genome-wide proportions within individuals sum to 1.0. We focus on the ancestry proportion (π) of one of the ancestries, so that the proportion of the unmodelled other ancestry is 1- π, and consider multiple admixed families with sibship size *n*. Define:

𝜋_𝑖_: True parental mean ancestry component of family *i*

𝜋_𝑖𝑗_: Ancestry of offspring *j* in family *i*

𝑑_𝑖𝑗_ = 𝜋_𝑖𝑗_ − 𝜋_𝑖_ : Deviation of offspring ancestry from the parental mean

𝜋^_𝑖_ = ∑^𝑛^ 𝜋_𝑖𝑗_ /𝑛 : Mean sibship ancestry component in family *i*_2_

By definition, 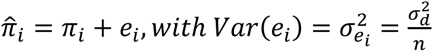

#### Generating and analysis models

Assume the following generating model:

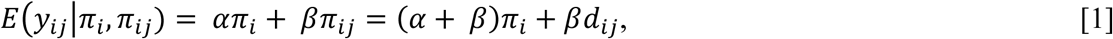

where α represents a between-family effect that is associated with ancestry and β represents the direct genetic ancestry effect. If 𝜋_𝑖_ is observed, then multiple linear regression of 𝑦_𝑖𝑗_ on 𝜋_𝑖_and 𝜋_𝑖𝑗_ will give unbiased estimates of parameters 𝛼 and 𝛽 because the model of analysis is the same as the assumed generating model. When 𝜋_𝑖_ is unobserved due to the unavailability of parental genotypes for offspring with phenotypic data, the sibship average ancestry proportion is used instead and the analysis model becomes: 𝑦_𝑖𝑗_ = 𝑏_𝐵_𝜋^_𝑖_ + 𝑏_𝑊_𝜋_𝑖𝑗_ + 𝜖_𝑖𝑗_

To derive the expectations of ordinary least-squares estimates of 𝑏_𝐵_ and 𝑏_𝑊_ we need the relevant variance and covariance terms:

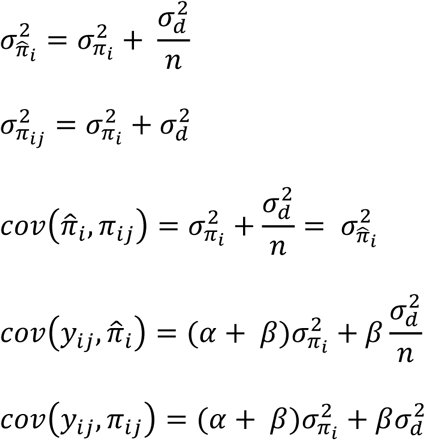

Solving for 𝑏_𝐵_ and 𝑏_𝑊_ using these (co)variance components gives:

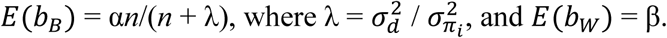

Hence, when parental genotypes are not observed, the estimated between-family effect is shrunk towards zero, depending on sibship size *n* and variance ratio λ. This shrinkage reflects the uncertainty of the true family mean ancestry component. The estimate of the direct effect remains unbiased^58^.

In the absence of family data, when the analysis is performed at the population level, the model simplifies to: 𝑦 = 𝑏_𝑃_𝜋 + 𝑒, resulting in the regression coefficient: 𝐸(𝑏_𝑃_) = β + α/(1 + λ). This expression demonstrates how the contribution of the between-family effect diminishes as λ increases, while the contribution of the direct genetic effect β remains consistent. These results allow us to predict the expected value of 𝑏_𝑃_ in a population sample from estimates of 𝑏_𝐵_ and 𝑏_𝑊_in family data,

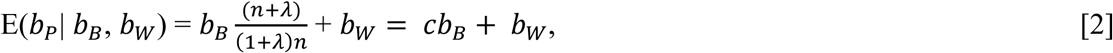

with sampling variance

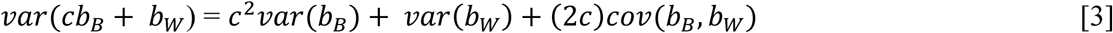

#### MCPS analyses

In MCPS data, there are four ancestries and for each individual the genome-wide proportions of ancestries sum to 1.0. There are only three estimable effects of ancestry and we modelled the effects of IAM, AFR and EAS, so that the results are relative to a EUR ancestry baseline. For the analysis of unrelated samples, let 𝜋_𝑗𝑘_be the genome-wide proportion of ancestry *k* of individual *j*. For each trait that we adjusted for age, age^2^ and sex and standardised to a mean of 0 and a variance of 1, we fitted the model

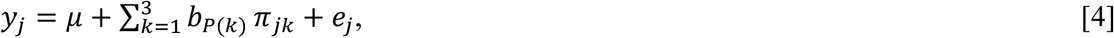

where 𝑏_𝑃(𝑘)_are the effects associated with ancestry. For the binary trait type 2 diabetes, the linear part of the model is the same but 𝑦_𝑗_ is replaced by logit(𝑃(𝑦_𝑗_ = 1)).

For the analysis of the family data, let 𝜋_𝑖𝑗𝑘_ be the genome-wide proportion of ancestry *k* of individual *j* in family *i*. For each standardised trait *y*, we fitted the model

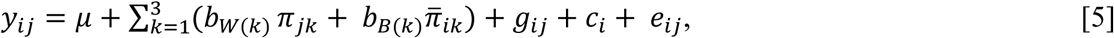

where 𝑏_𝑊(𝑘)_and 𝑏_𝐵(𝑘)_are the within-family direct and between-family indirect effect of ancestry *k*, 𝜋̅_𝑖𝑘_is the average genome-wide proportion of ancestry *k* in family *i*, 𝑔_𝑖𝑗_is the genetic value of individual *j* in family *i* and 𝑐_𝑗_ is a family effect for family *i*. For 𝜋̅_𝑖𝑘_ we took the average parental estimates if available and otherwise used the sibship average ancestry proportion. Sibling covariance for individual *j* and *l* in family *i* was modelled as,

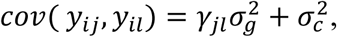

with 𝛾_𝑗𝑙_the proportion of the genome shared identical-by-descent. For the generalised linear mixed model, 𝑦_𝑖𝑗_is replaced by logit(𝑃(𝑦_𝑖𝑗_ = 1)) and only a single random family effect 𝑐_𝑖_was fitted to account of the covariance between siblings.

In addition to estimation the population effects 𝑏_𝑃(𝑘)_ directly in the unrelated set, we also predicted their expected value from the estimates of 𝑏_𝐵(𝑘)_and 𝑏_𝑊(𝑘)_obtained from the linear mixed model for each ancestry, using λ values from Extended Data Table 3 and the average sibship size for *n*. Standard errors of the predicted values were calculated from Eq[3], using the sampling (co)variance of the regression coefficients from the option *--reml-est-fix-varcov* in GCTA^52^.

Note that in the MCPS data, the within-family ancestry segregation variance 𝜎_d_^2^ is very small relative to the between-family variance 𝜎^2^ (Extended Data Table 3), so that 𝜆 ≈ 0, 𝐸(𝑏_𝐵_) ≈ 𝛼 and 𝐸(𝑏_𝑃_) ≈ 𝛼 + 𝛽, the sum of the between and within-family effects.

## Supplementary Figures

**Supplementary Figure 1:**
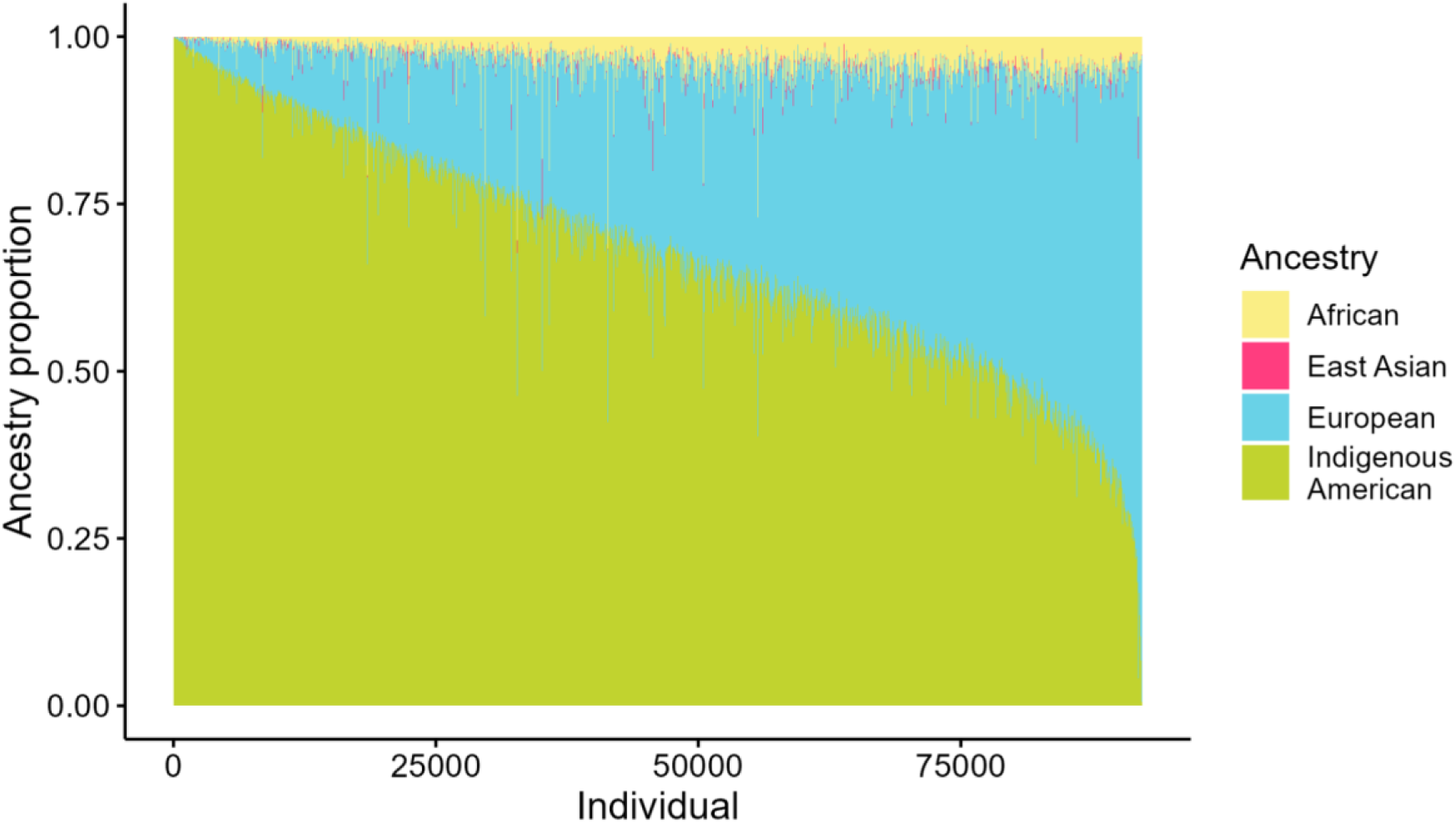
**Ancestry proportion distribution of the 92,297 study individuals.**

**Supplementary Figure 2:**
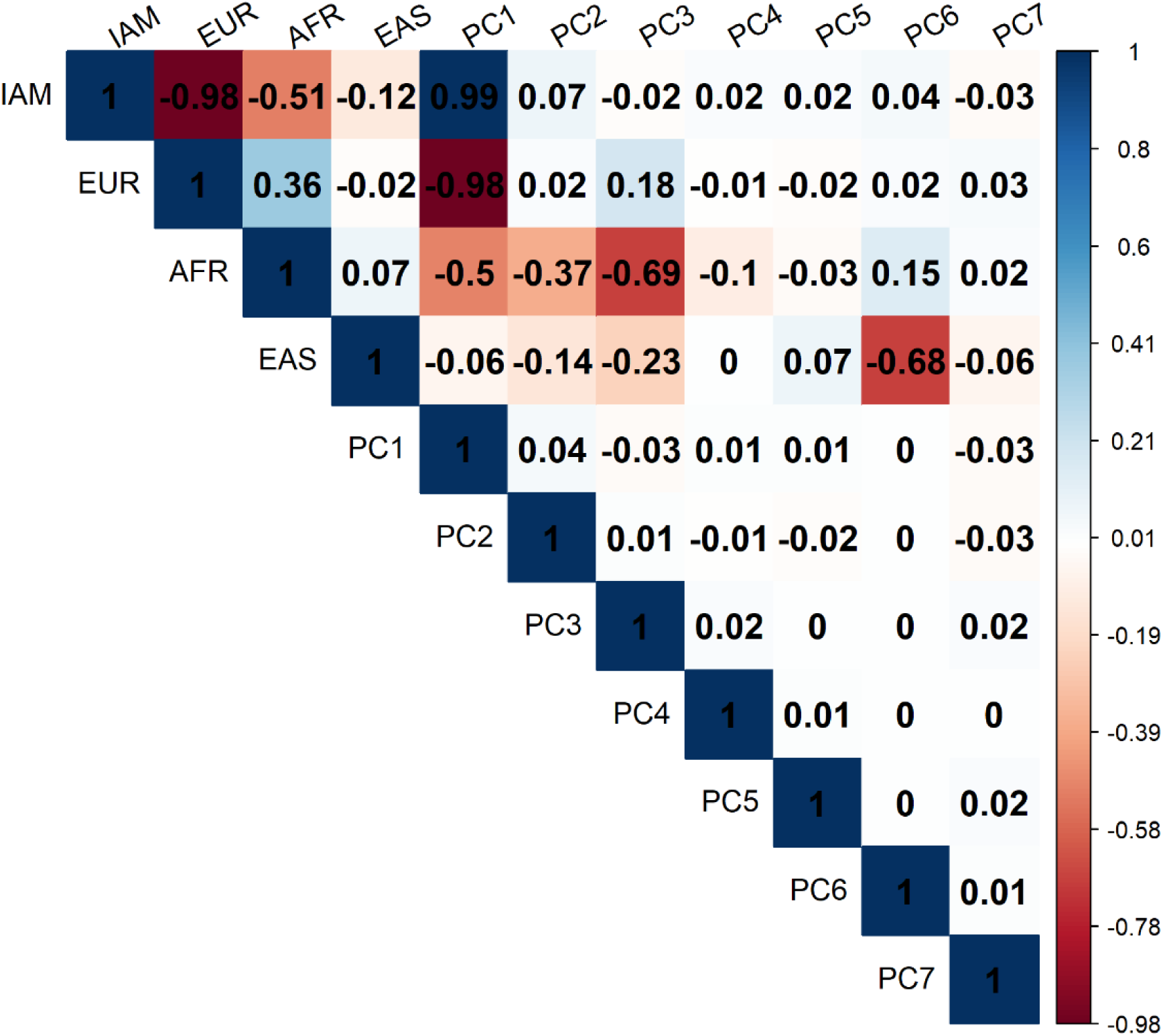
**Correlation plot of ancestry proportions and PCs in a sample of 52,583 unrelated individuals.**

**Supplementary Figure 3:**
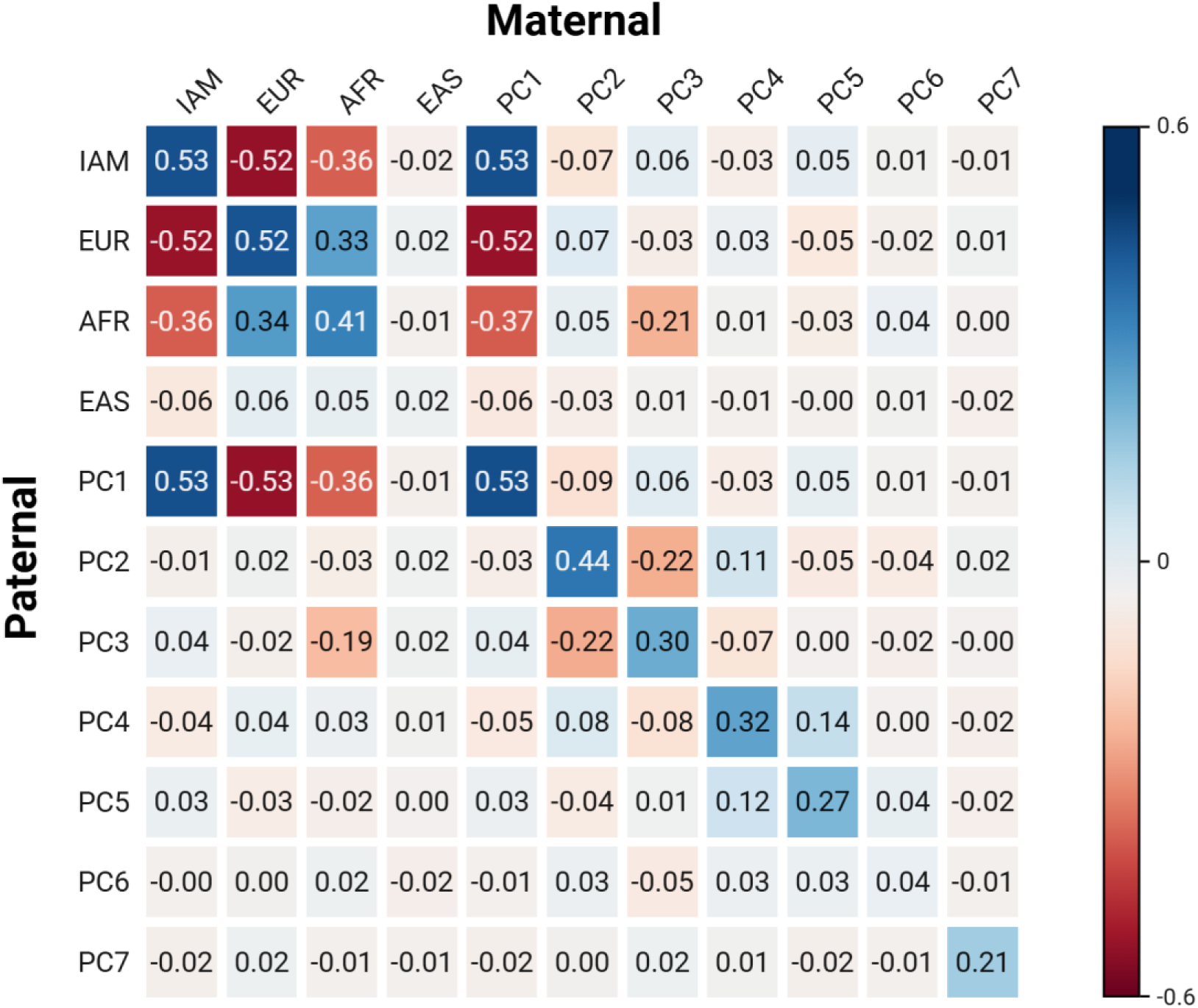
**Correlations between paternal and maternal ancestry proportions and PCs observed from 2,847 parent pairs.**

**Supplementary Figure 4:**
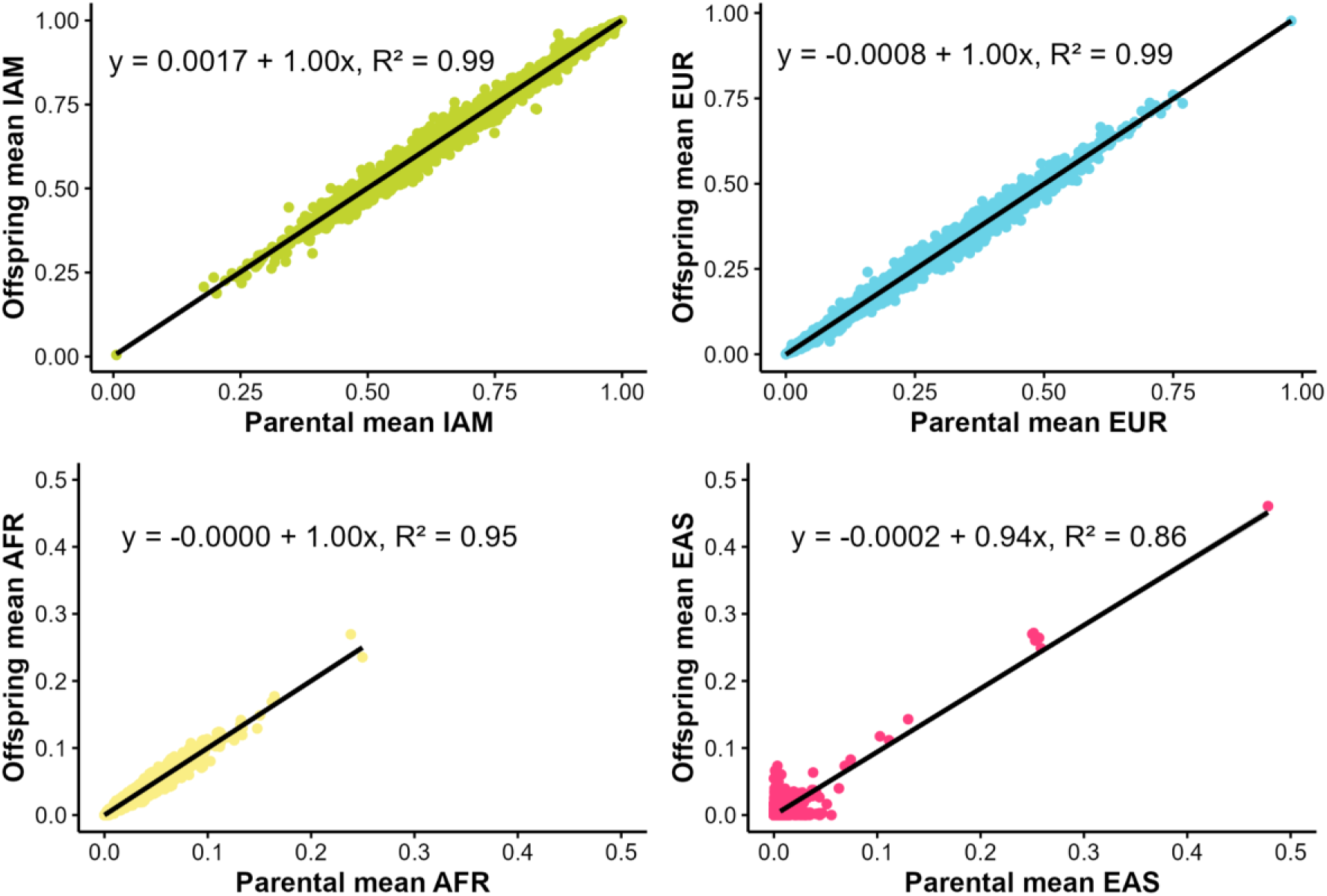
**Regression of offspring average ancestry proportion on the parental average ancestry proportion.** Correlations were observed in families (*n* = 2,847) with available paternal and maternal genotype data.

**Supplementary Figure 5:**
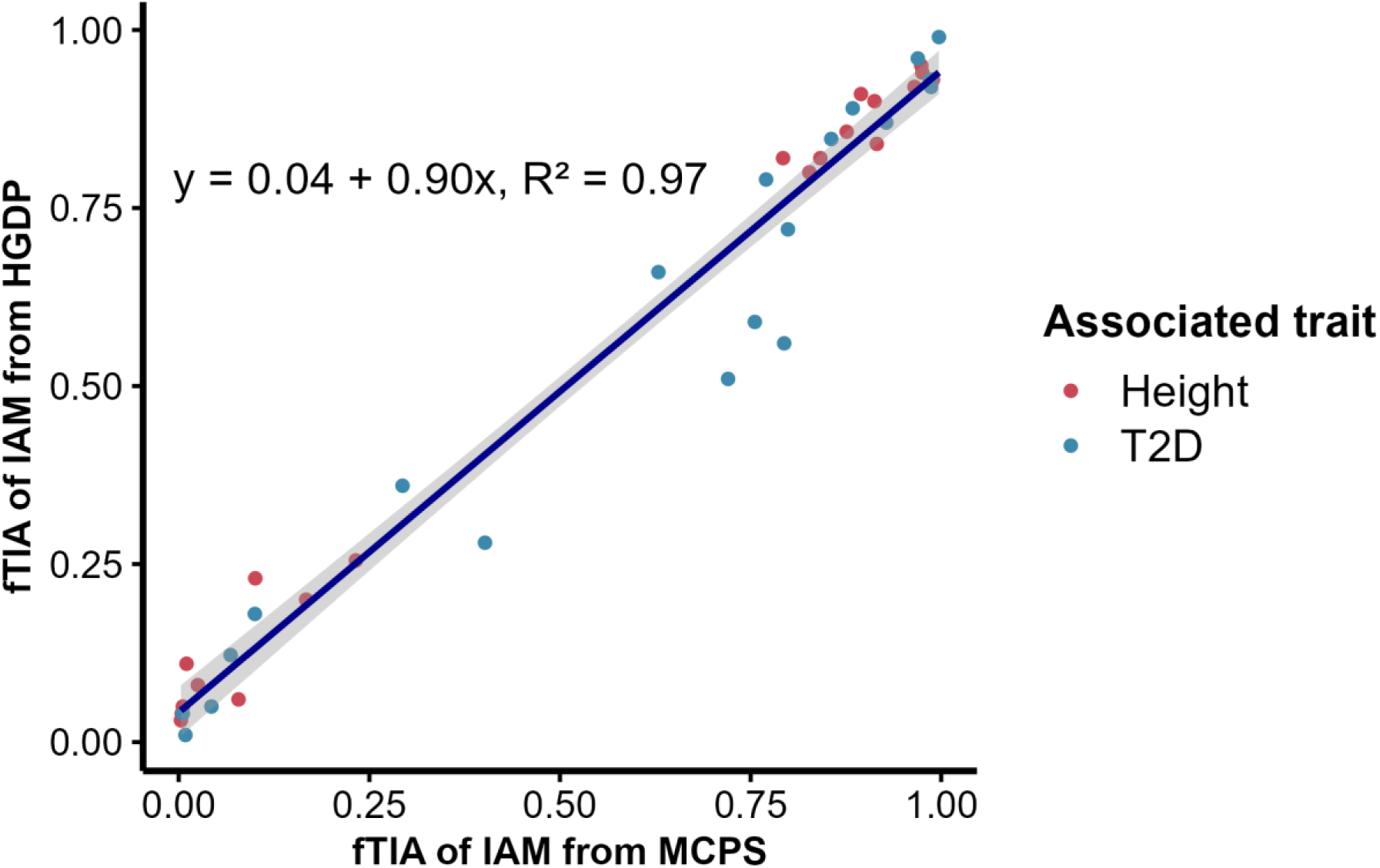
**Comparison of IAM allele frequencies in MCPS and HGDP at the top 20 loci with the largest EUR–IAM differences for height and type 2 diabetes separately.** Abbreviation: fTIA = frequency of trait-increasing alleles, IAM = Indigenous American, EUR = European.

**Supplementary Figure 6:**
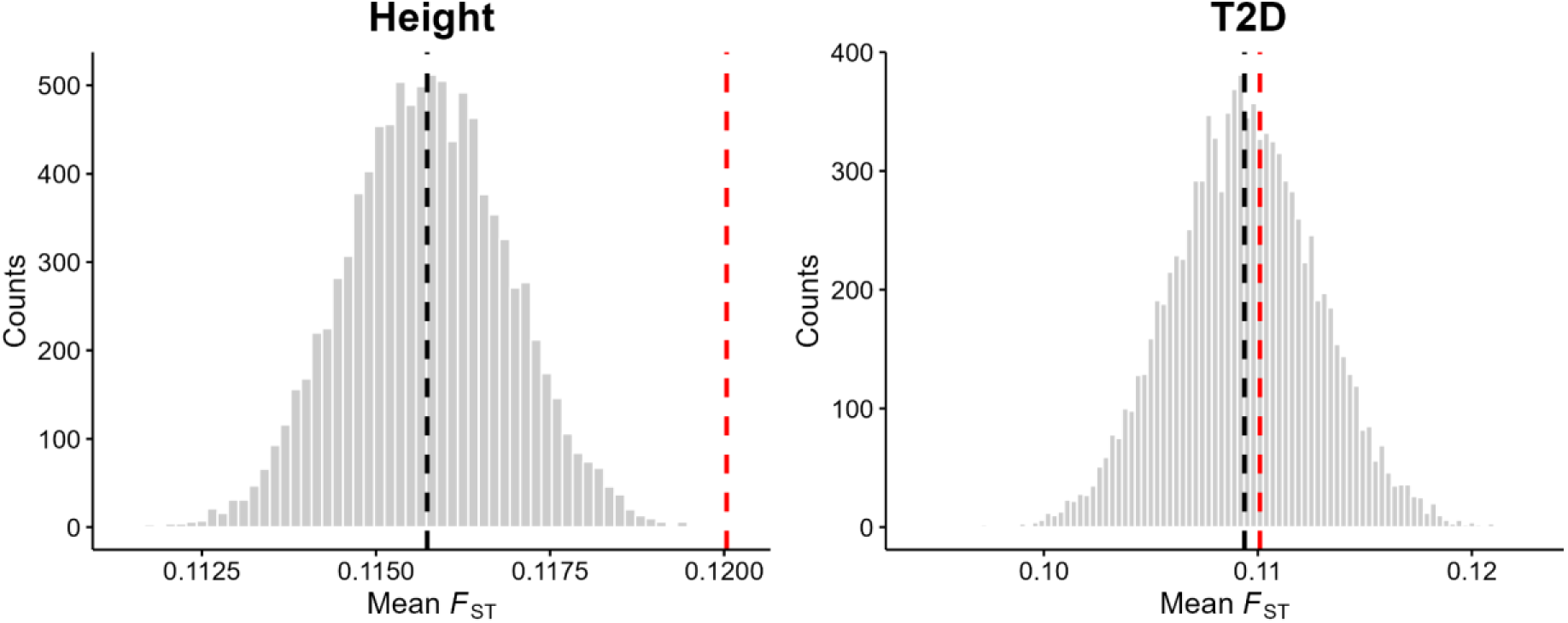
**Comparison of mean *F*_ST_ values between IAM and EUR for height and type 2 diabetes.** Histogram showing the null distribution of mean *F*_ST_ values derived from control SNP sets for height and type 2 diabetes, and the black dashed line indicates the mean value of the distribution. The red dashed line indicates the observed mean *F*_ST_ of the trait-associated SNPs.

**Supplementary Figure 7:**
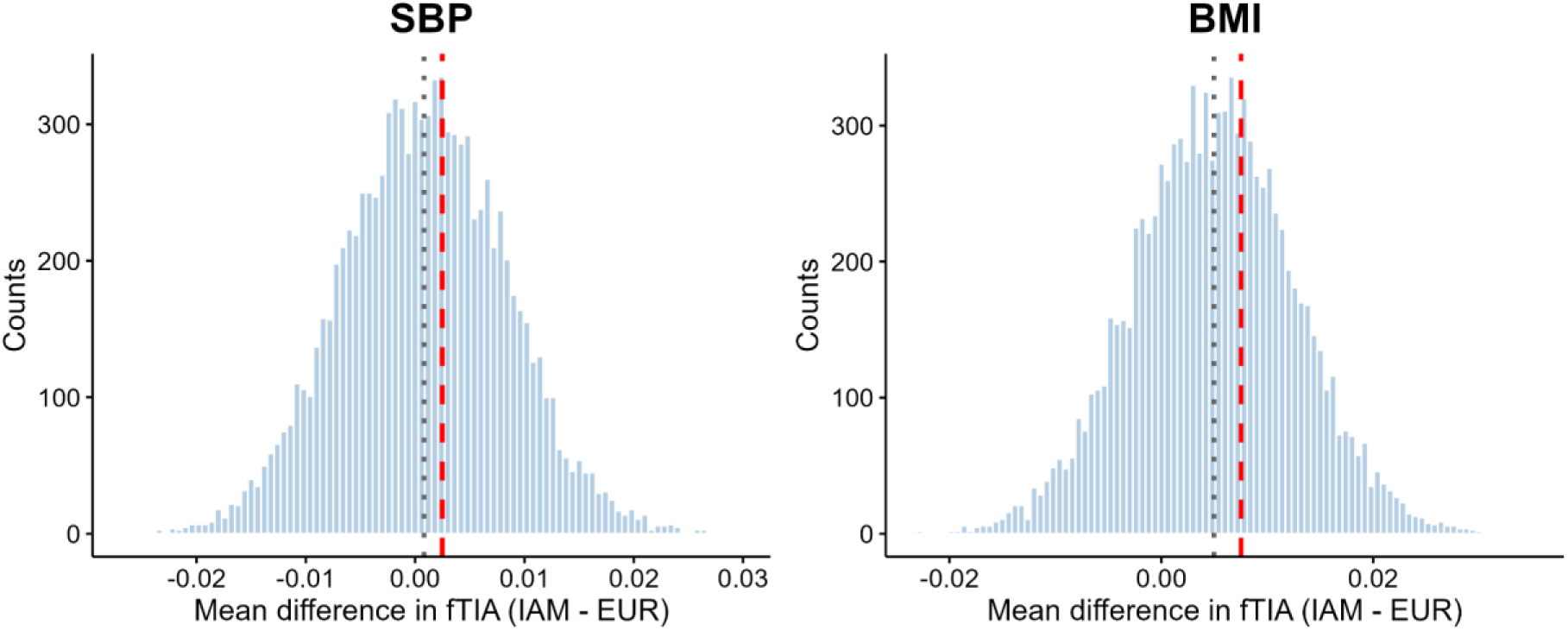
**Mean difference in the frequency of trait-increasing alleles between IAM and EUR populations for SBP and BMI.** The histogram shows the distribution of fTIA differences for control SNPs, and the grey dashed line represents its mean value. The red dashed line indicates the mean value of the fTIA difference for the trait-associated SNPs.

**Supplementary Figure 8:**
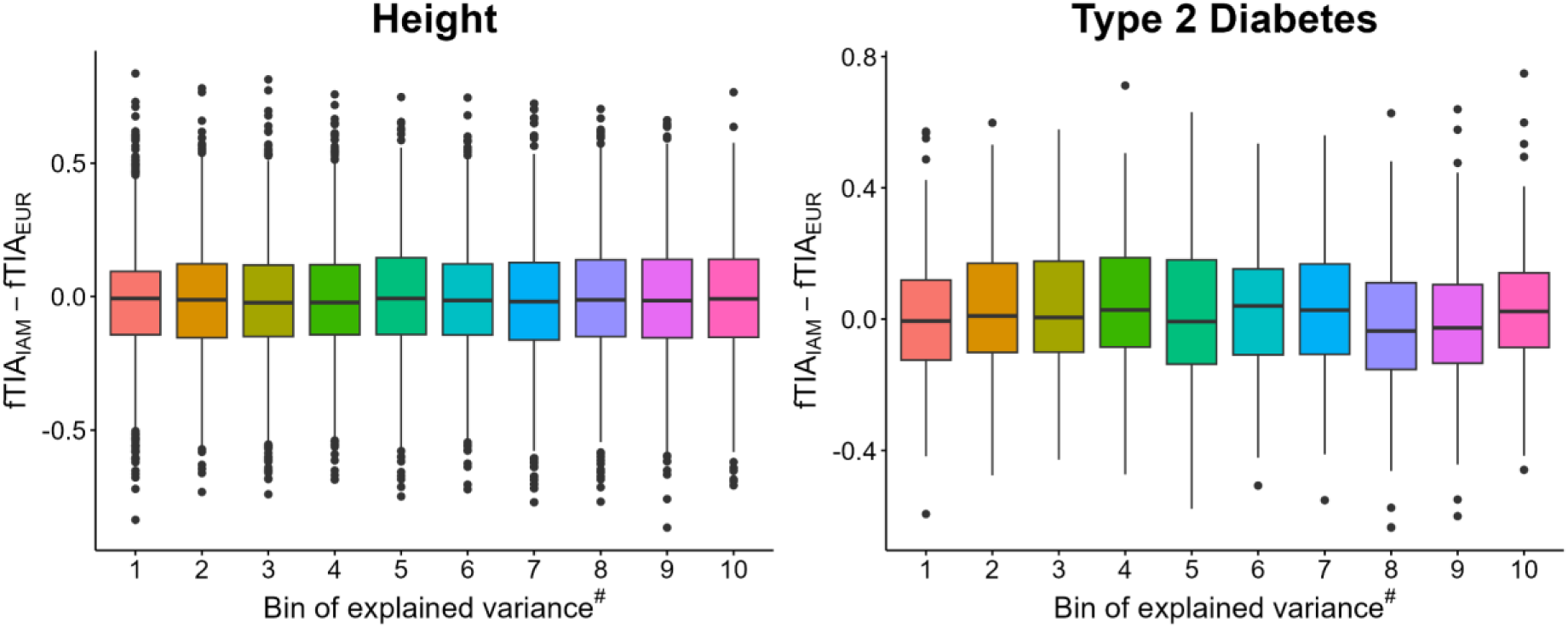
**fTIA differences against variance explained for 11,778 and 1,208 genome-wide significant variants of height and T2D, respectively.** The x-axis is the predicted variance^#^ explained in EUR, assuming Hardy-Weinberg proportions of genotypes. The y-axis is the difference in trait-increasing allele frequency between IAM and EUR. ^#^Explained Variance = 2 × fTIA_EUR_ × (1-fTIA_EUR_) × β^2^, where β is the estimated effect size from the discovery GWAS and fTIA_EUR_ the frequency of trait-increasing alleles in the UK Biobank. Loci were grouped into 10 variance bins, with an equal number of loci per bin. For both height and T2D, there is no statistical association between increasing trait differentiation and predicted variance explained.

**Supplementary Figure 9:**
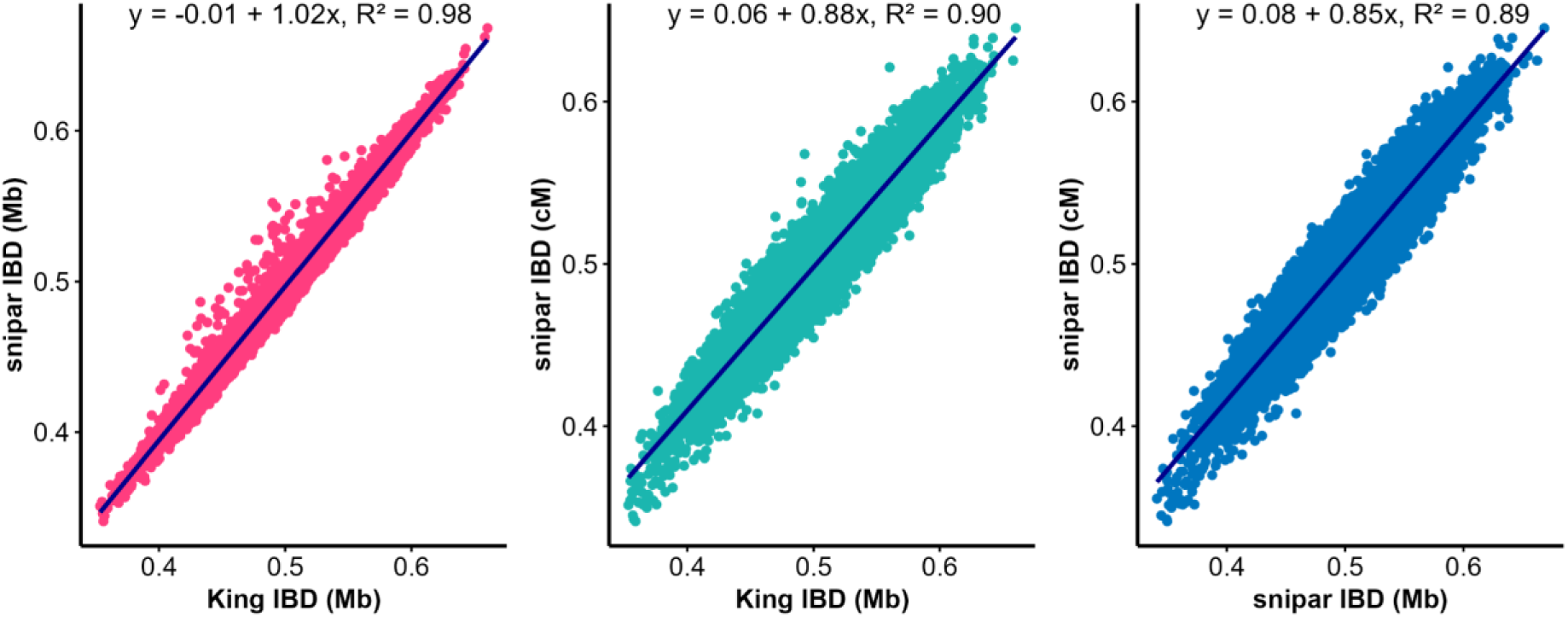
**Comparison between genome-wide IBD proportions calculated by KING and snipar from 30,407 full-sibling pairs.** Abbreviation: IBD = identical-by-descent, cM = centimorgan.

## Notes

### Competing Interest Statement

The authors have declared no competing interest.

### Author Declarations

Ethics approval was obtained from the Mexican Ministry of Health, the Mexican National Council for Science and Technology, and the University of Oxford, UK.

